# Renal Impairment and Parkinson’s Disease in Cardiovascular Patients: Associations, Pre-diagnostic Trajectories, and Predictive Enhancement

**DOI:** 10.64898/2026.04.07.26350284

**Authors:** Jike Qi, Ping Zeng

## Abstract

**Background:** Renal impairment is associated with increased risk of Parkinson’s disease (PD) in general populations; however, the renal-PD link within cardiovascular disease (CVD) patients remains unclear through the high comorbidity of renal dysfunction and elevated PD risk among this special population.

**Objectives:** To assess renal function’s association, longitudinal trajectories and predictive value for PD specifically within a cardiovascular disease cohort.

**Methods:** Among 29,266 UK Biobank CVD patients, we assessed baseline renal function via creatinine-based (eGFRcr) and cystatin C-based (eGFRcys) estimated glomerular filtration. Multivariable Cox regression analyzed associations with incident PD and all-cause mortality, with wide sensitivity analyses addressing reverse causation/confounding. Nested case-control analysis characterized pre-PD eGFR trajectories over 14 years. We finally evaluated whether renal function improved the PREDICT-PD’s predictive ability.

**Results:** Over a median 13.1-year follow-up, 489 incident PD cases and 5,919 deaths occurred. Lower eGFR levels exhibited dose-dependent associations with increased PD risk (eGFRcr: HR=0.87 [0.80∼0.95]; eGFRcys: HR=0.90 [0.82∼0.99]) and all-cause mortality (eGFRcr: HR=0.77 [0.75∼0.79]; eGFRcys: HR=0.64 [0.63∼0.66]). Pre-PD eGFR trajectories diverged significantly from controls starting over 14 years before diagnosis. eGFR-defined chronic kidney disease (<60 ml/min/1.73m^2^) conferred 38∼60% higher PD risk and 159∼234% elevated mortality risk, and could significantly enhance PREDICT-PD’s discrimination, with a 1.18∼1.34% increase in prediction accuracy.

**Conclusions:** Impaired renal function is an independent PD and all-cause mortality risk factor of CVD patients, preceded by a slow, progressive eGFR decline starting >14 years before diagnosis. Incorporating renal function substantially improves PD risk prediction in this population.

## Introduction

Parkinson’s disease (PD) is among the most prevalent and severe neurodegenerative disorders, affecting more than 12 million people worldwide and imposing profound human and economic burdens 1–3. Population-level evidence has uncovered a compelling association of renal impairment with increased PD risk and progression, with chronic kidney disease (CKD) conferring a 20% elevated PD risk 4 and acute kidney injury related to a 47% higher risk 5. A recent UK Biobank analysis quantified this link in the general population, reporting a 1.9-fold elevation in PD incidence among individuals with severely reduced renal function (estimated glomerular filtration rate [eGFR] <30 ml/min/1.73m^2^) 6.

One of the central mechanisms implicated in the renal-PD connection involves cardiorenal crosstalk 7. Cardiorenal crosstalk, manifesting as impaired vascular health, endothelial dysfunction, and neurohormonal activation, can drive mitochondrial dysfunction and oxidative stress 7, which may promote α-synuclein aggregation and neurodegeneration — hallmark processes in PD 8. Importantly, this cardiorenal interplay is most pronounced in the context of established cardiovascular disease (CVD) 9. Consequently, investigating renal function and PD risk specifically within the CVD population is essential for elucidating the role of this critical pathway in PD pathogenesis, addressing a substantial gap in current knowledge.

Focusing on CVD patients holds particular clinical significance. This population bears a disproportionately high burden of both renal impairment, affecting 42∼63% with moderate-to-severe renal dysfunction 10, and neurodegenerative disorders, including a 55% elevated PD risk compared with individuals without CVD 11. This convergence of cardiovascular, renal, and neurological pathologies compounds morbidity, mortality, and healthcare expenditures 12. Understanding how renal dysfunction modulates PD susceptibility in CVD patients is therefore pivotal for identifying modifiable risk factors and developing strategies to protect neurological health in this large and vulnerable group. However, no prior studies have explicitly examined the relationship between renal impairment and PD risk confined to individuals with pre-existing CVD.

Additionally, significant methodological limitations constrain the validity of current renal-PD research. For example, residual confounding 13,14 remains inadequately addressed, potentially obscuring true causal pathways. Concurrently, unmanaged competing mortality risks 15 introduce substantial selection bias: severe CVD events in this comorbid population may preclude PD ascertainment, systematically excluding high-risk individuals from observation and distorting association estimates. Furthermore, the absence of long-term repeated measures data has impeded the characterization of renal function trajectories leading up to PD onset, hindering the identification of optimal screening windows 16. Finally, despite renal function markers (e.g., eGFR) being readily available within CVD care settings, existing mainstream PD prediction models largely neglect to incorporate them 17.

To address these knowledge gaps, we here utilized the prospective UK Biobank cohort to construct a multidimensional analytical framework. Within the CVD subcohort, we examined the associations of two mutually validated renal function metrics (eGFRcr and eGFRcys) with incident PD and all-cause mortality, with the latter serving as a composite indicator of renal impairment burden. Triple-robust estimation targeting confounding and cause-specific risk regression was used to performing sensitivity analyses. We then conducted nested case-control analysis to characterize the 14-year longitudinal eGFR trajectories preceding PD diagnosis or mortality, respectively. We also explicitly evaluated whether incorporating renal function metrics could enhance the predictive performance of the PREDICT-PD model for risk prediction 17. Figure 1 presents a schematic overview of the analytical framework.

**Figure 1.**
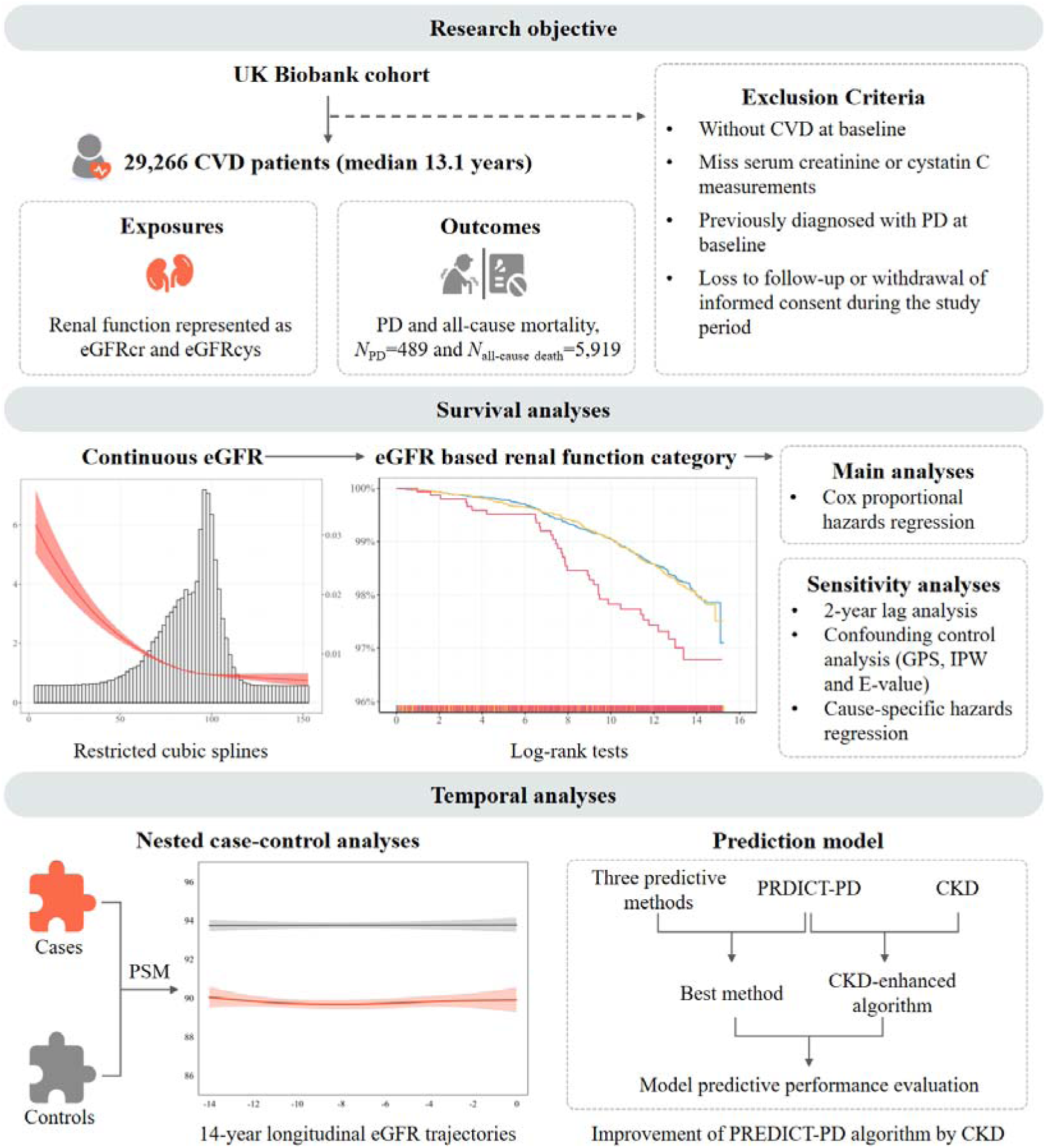
Flow chart of the study. Two eGFR-related traits serve as the exposures, with PD and all-cause mortality as the outcomes. We investigated the phenotypic relation between the two. CVD, cardiovascular disease; eGFR, estimated glomerular filtration rate; eGFRcr, estimated glomerular filtration rate based on creatinine; eGFRcys, estimated glomerular filtration rate based on cystatin C; PD, Parkinson’s disease; GPS, generalized propensity score; IPW, inverse probability weight; PSM, propensity score matching.

## Materials and methods

### Data sources and variable definition

#### UK Biobank cohort

Our analysis focused on the subcohort of the UK Biobank 18 with prevalent CVD, mainly composed of coronary heart disease and stroke 19 (Table S1). To ensure homogeneity and adequate statistical power, we restricted this subpopulation to participants of European ancestry, and excluded individuals with the following characteristics: (i) missing serum creatinine or cystatin C measurements; (ii) previously diagnosed with PD at baseline; (iii) loss to follow-up or withdrawal of informed consent during the study period. Ultimately, a total of 29,266 participants with CVD were included.

#### Definition of exposures and outcomes

eGFR was calculated using two validated CKD-EPI equations for eGFRcr and eGFRcys 20,21, with lower eGFR indicating impaired renal function (Supplementary File). In subsequent analyses, eGFR was standardized to improve model interpretability. The primary outcome was incident PD, defined as the first recorded diagnosis of PD during follow-up. PD cases were identified through structured electronic health records (EHRs) using the ICD-10 codes (Table S1). The secondary outcome was all-cause mortality, and mortality data were obtained from national death registries linked to EHRs. The follow-up period spanned from the baseline assessment date (study enrollment) to the earliest of PD diagnosis, all-cause death, or the administrative censoring date (July 19, 2022).

#### Ascertainment of covariates

We selected the following baseline covariates (Table S1): socioeconomic factors (Thompson deprivation index [TDI], educational level, and income), behavioral factors (alcohol consumption status, smoking status, physical activity status, and healthy diet score), health status (body mass index [BMI], type 2 diabetes, hypertension, dyslipidemia), top-frequency CVD medication use (angiotensin-converting enzyme inhibitors, angiotensin II receptor blockers, β-blockers, calcium channel blockers, diuretics, non-steroidal and anti-inflammatory drugs) 22,23 (Table S2), family history of PD and top ten genetic principal components. Age and gender were inherently adjusted in eGFR calculations and thus excluded from multivariable models to avoid over-adjustment. Multiple imputation by chained equations was used to estimate missing values for each covariate 24.

### Statistical analyses

#### Survival analyses evaluating the relation between renal function and PD

Our study adhered to the STROBE guideline 25, with the completed checklists shown in Table S3. Baseline characteristics were stratified by incident PD status during follow-up. Quantitative variables were expressed as mean ± standard deviation (SD), and categorical variables as frequency (proportion). Differences between groups were tested using Student’s *t*-test for continuous variables and Pearson χ^2^ test for categorical variables.

To evaluate the associations of baseline eGFR with incident PD and all-cause mortality, we implemented mutually confirmatory analytic approaches. Continuous eGFR was modeled with restricted cubic splines (with knots placed at the 10^th^, 50^th^, and 90^th^ percentiles) to examine non-linear relationships. Participants were then categorized into three renal function groups according to KDIGO guideline 21: normal (eGFR≥90 ml/min/1.73m^2^), mildly impaired (60 ml/min/1.73m^2^ eGFR<90 ml/min/1.73m^2^), and CKD (eGFR<60 ml/min/1.73m^2^). We calculated mean time-to-PD-onset and mean survival time for each group while accounting for baseline age, and constructed Kaplan-Meier curves, with between-group differences assessed via log-rank tests. Multivariable Cox proportional hazards models were employed, adjusting for covariates described earlier. The proportional hazards assumption was verified through Schoenfeld residual testing (global *P*>0.05) 26. Results were presented as hazard ratios (HRs) with 95% confidence intervals (CIs).

#### Assessment of temporal stability, balance, weights, and hidden biases

Three sensitivity analyses were conducted to assess the robustness of the observed associations. First, to mitigate potential reverse causation bias where preclinical PD could influence eGFR, we excluded participants diagnosed with PD or deceased within the initial two years of follow-up. Second, to balance baseline confounders across continuous eGFR levels, we modeled generalized propensity score (GPS) for eGFR conditional on covariates 13. Deriving inverse probability weight (IPW) as the inverse of each participant’s observed eGFR probability density 14, we applied these weights directly to Cox regression for incident PD and all-cause mortality. This approach constructed a pseudo-population with balanced covariates, enabling subsequent calculation of E-values to quantify the strength of unmeasured confounding needed to nullify associations 14. Third, to address potential bias from competing mortality risk, where death from severe CVD could preclude PD detection, we implemented cause-specific hazards regression for incident PD 15, explicitly censoring all non-PD deaths.

#### Nested case-control study assessing time-dependent eGFR shifts preceding PD and all-cause mortality

To map dynamic trajectories of renal function preceding PD diagnosis or preceding all-cause mortality, we conducted a nested case-control study. To this aim, we identified incident cases during follow-up and performed 1:1 PSM using covariates 27. Given that age and sex were already adjusted when calculating eGFR, we did not include them in the matching. With cases matched to controls remaining PD-free or death-free until the case’s diagnosis date, and decedents matched to surviving controls, each case defined an observation window from baseline to their event date, with matched controls assigned exactly the same baseline-to-event-duration period. Within this framework, eGFR measurements during the 14-year pre-event period (-14 to 0 years) were analyzed. Dynamic trajectories were modeled with locally weighted scatterplot smoothing, enabling direct comparison of pre-outcome trends within temporally aligned case-control pairs.

#### Predictive value of renal function on the risk of incident PD

Using our cohort, we externally validated the PREDICT-PD algorithm 17 for predicting incident PD. This algorithm calculates the baseline predicted probability as *P*_0_=*r*_0_/(1+*r*_0_), where *r*_0_ represents the PD odds derived from established independent risk factors (Supplementary File). To evaluate *P*_0_’s predictive performance, we carried out Monte Carlo cross-validation (200 iterations via 70%:30% training-validation splits) 28. Within each training set, *P*_0_ served as the input predictor for three distinct models, including logistic regression, XGBoost 29, and LightGBM 30, trained to predict incident PD. Performance was assessed in each corresponding validation set using the area under the receiver operating characteristic curve (AUC), identifying the best-performing model across all iterations.

To assess renal function’s incremental value, with eGFR-defined CKD (eGFR<60 ml/min/1.73m²) as our proxy variate, we enhanced the baseline PREDICT-PD odds within each training set. Specifically, we fit a logistic regression for incident PD, adjusting for *P*_0_ and external covariates to obtain the adjusted relative risk (RR_CKD_) associated with CKD. This RR_CKD_ was then used to augment *r*_0_: participants with CKD were assigned *r*_1_=RR_CKD_×*r*_0_, while those without retained *r*_1_=*r*_0_. We subsequently computed the CKD-enhanced probability (*P*_1_=*r*_1_/[1+*r*_1_]) 31. Finally, utilizing predictions from the previously identified best-performing model in each validation cohort, we compared the predictive performance of *P*_1_ against the original *P*_0_ using AUCs’ difference ratio, net reclassification improvement index (NRI) and integrated discrimination improvement index (IDI). All metrics were aggregated as means with 95%CIs across all iterations.

### Statistical software and significance criteria

All statistical analyses were performed using R v4.5.0. The association test was two-sided, with *P*<0.05 considered statistically significant.

## Results

### Baseline characteristics

We analyzed 29,266 CVD patients (33.4% female) with a mean age of 62.2 years (SD=6.1) (Table 1). During a median follow-up of 13.1 years (interquartile range 12.2∼14.0 years), we discovered that this subpopulation exhibited markedly higher PD incidence (1.7%=489/29,266) and all-cause mortality (20.2%=5,919/29,266) compared with the general European-ancestry population in the UK Biobank (0.7%=3,211/440,592 for incidence PD and 7.6%=33,300/440,592 for mortality).

**Table 1.**
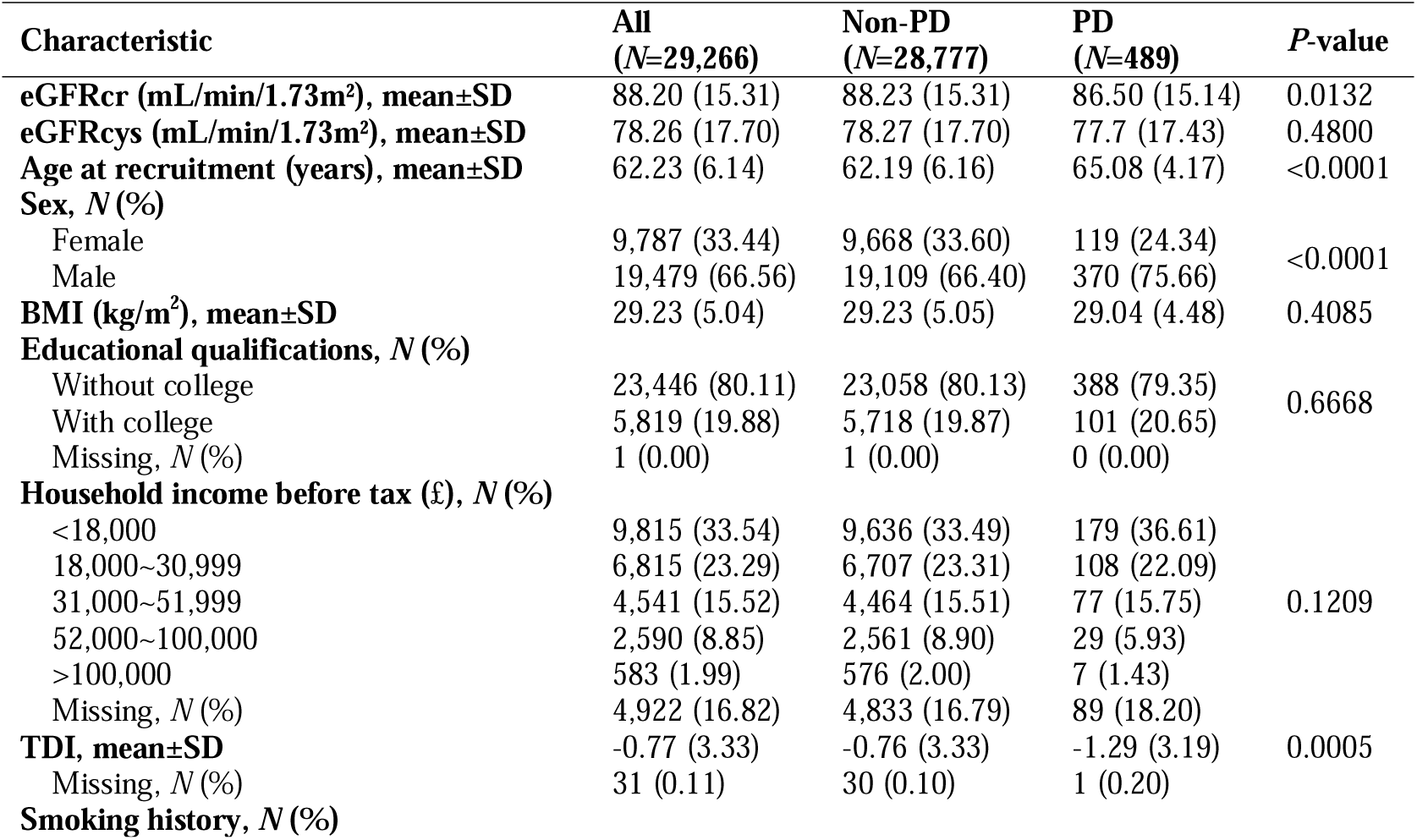
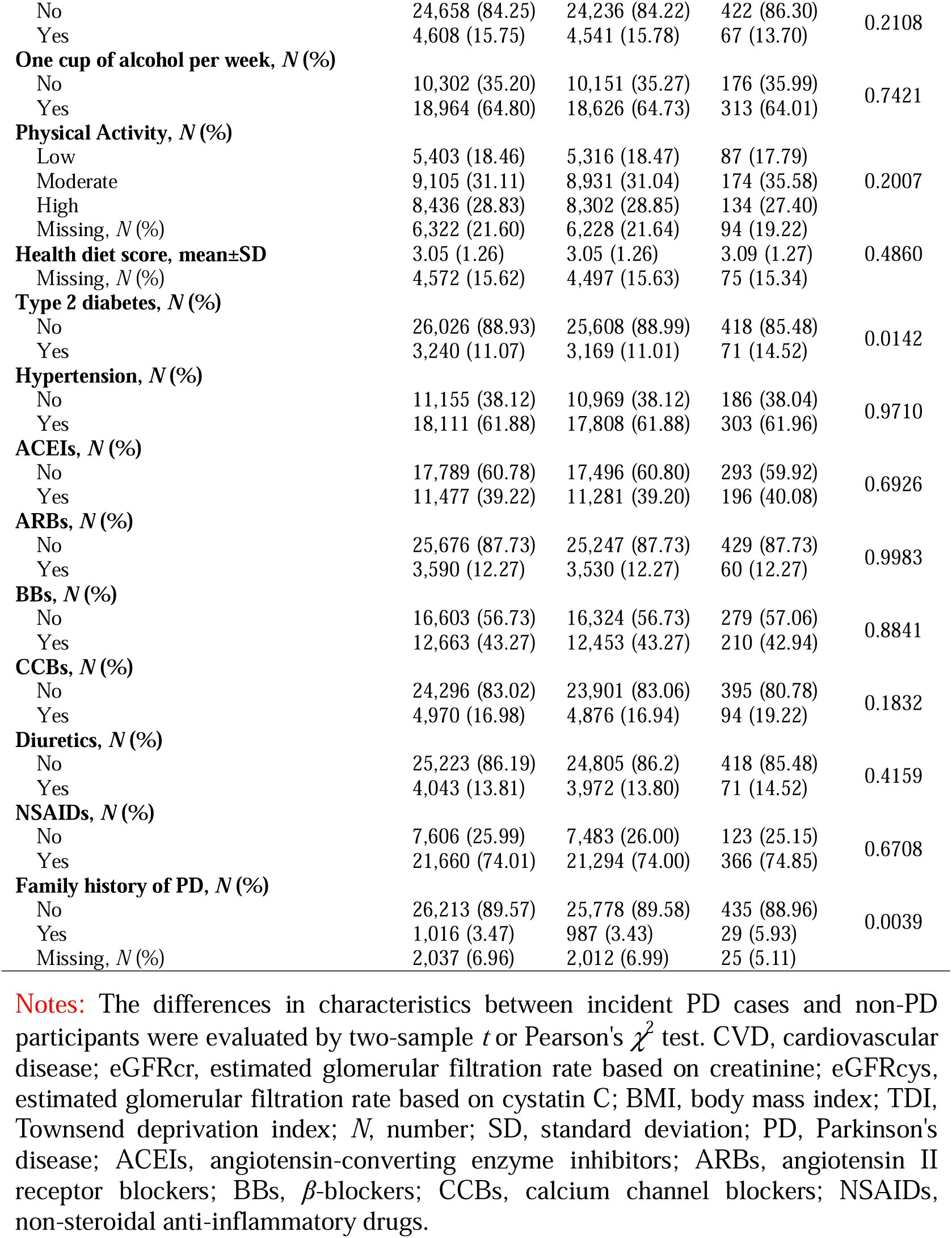
Baseline characteristics of all included participants in CVD population.

Compared with non-PD individuals, PD patients were older (65.1 vs. 62.2 years) and were more likely to be male (75.7% vs. 66.4%). Regarding renal function, eGFRcr was lower in the PD group (86.5 vs. 88.2), while eGFRcys was comparable between the two groups. Socioeconomic status, as measured by the TDI, was smaller in the PD group (-1.1 vs. -1.5). The PD group showed a higher prevalence of type 2 diabetes (14.5% vs. 11.0%) and family history of PD (5.9% vs. 3.4%). No significant between-group differences were observed in BMI, educational qualifications, household income, smoking or alcohol consumption history, physical activity, healthy diet scores, and any CVD medication use.

### Association of renal function with PD and all-cause mortality

The restricted cubic spline analysis revealed largely monotonic relationships between either eGFRcr or eGFRcys and incident PD (Figure 2A-B), with non-linear (L-shaped) associations observed for all-cause mortality (Figure 2C-D). In multivariable-adjusted Cox models, each 1-SD reduction in eGFRcr or eGFRcys was related to a 12.7% (4.7∼20.0%) or 10.1% (1.2∼18.2%) increase in PD risk (Table 2), respectively. The inverse relationship was markedly stronger for all-cause mortality, with each 1-SD decrease in eGFRcr or eGFRcys conferring a 22.8% (21.1∼24.6%) or 35.9% (34.3∼37.5%) higher mortality risk.

**Figure 2.**
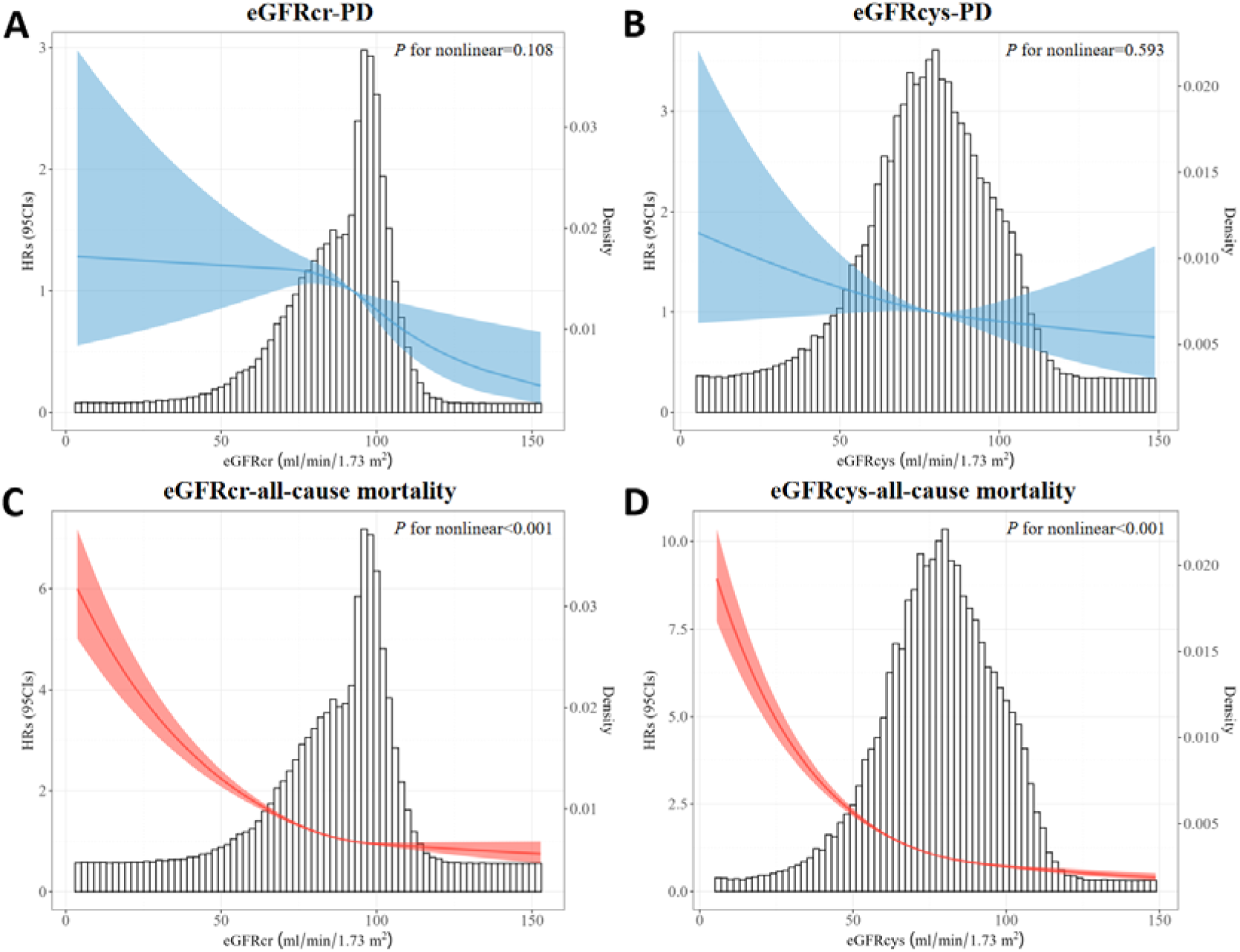
(A) Restricted cubic spline curve reflecting the relation of eGFRcr with PD. (B) Restricted cubic spline curve reflecting the relation of eGFRcys with PD. (C) Restricted cubic spline curve reflecting the relation of eGFRcr with all-cause mortality. (D) Restricted cubic spline curve reflecting the relation of eGFRcys with all-cause mortality. To visually demonstrate the biological significance of the results, we use the original values of eGFR here. PD, Parkinson’s disease; eGFRcr, estimated glomerular filtration rate based on creatinine; eGFRcys, estimated glomerular filtration rate based on cystatin C; eGFR, estimated glomerular filtration rate; CKD, chronic kidney disease; HRs, hazard ratios; CIs, confidence intervals.

**Table 2.**
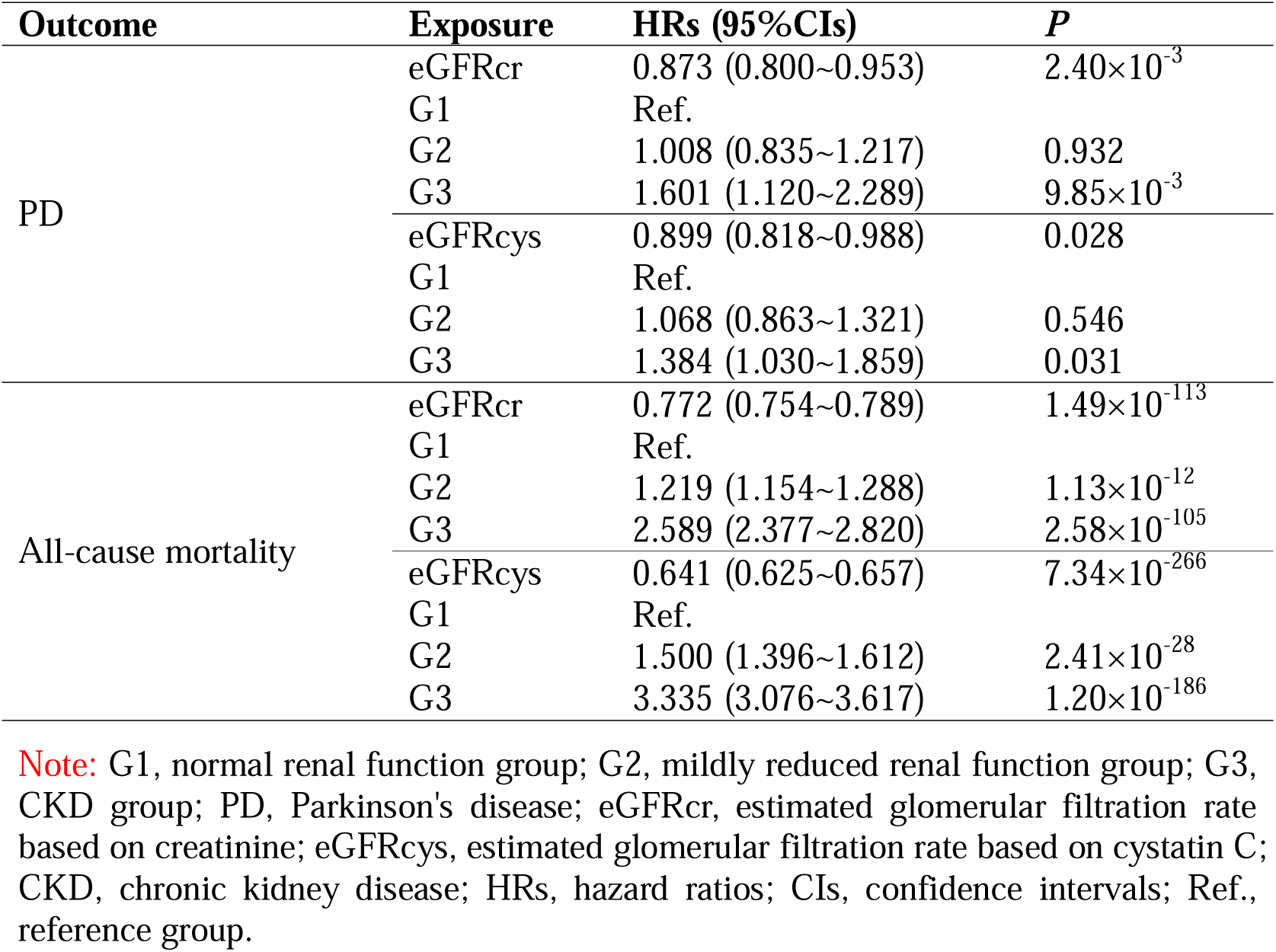
Phenotypic association of renal function with PD.

| Outcome | Exposure | HRs (95% CIs) | P |
| --- | --- | --- | --- |
| PD | eGFRcr | 0.873 (0.800~0.953) | $2.40 \times 10^{-3}$ |
|  | G1 | Ref. |  |
|  | G2 | 1.008 (0.835~1.217) | 0.932 |
| | G3 | 1.601 (1.120~2.289) | $9.85 \times 10^{-3}$ |
|  | eGFRcys | 0.899 (0.818~0.988) | 0.028 |
|  | G1 | Ref. |  |
|  | G2 | 1.068 (0.863~1.321) | 0.546 |
|  | G3 | 1.384 (1.030~1.859) | 0.031 |
| All-cause mortality | eGFRcr | 0.772 (0.754~0.789) | $1.49 \times 10^{-113}$ |
|  | G1 | Ref. |  |
| | G2 | 1.219 (1.154~1.288) | $1.13 \times 10^{-12}$ |
| | G3 | 2.589 (2.377~2.820) | $2.58 \times 10^{-105}$ |
| | eGFRcys | 0.641 (0.625~0.657) | $7.34 \times 10^{-266}$ |
|  | G1 | Ref. |  |
| | G2 | 1.500 (1.396~1.612) | $2.41 \times 10^{-28}$ |
| | G3 | 3.335 (3.076~3.617) | $1.20 \times 10^{-186}$ |

**Note:** G1, normal renal function group; G2, mildly reduced renal function group; G3, CKD group; PD, Parkinson's disease; eGFRcr, estimated glomerular filtration rate based on creatinine; eGFRcys, estimated glomerular filtration rate based on cystatin C; CKD, chronic kidney disease; HRs, hazard ratios; CIs, confidence intervals; Ref., reference group.

When eGFR was classified, initial Kaplan-Meier curves in Figure 3A-D visually demonstrated that worsening renal function (categorized by both eGFRcr and eGFRcys levels) correlated with progressively PD events and reduced overall survival probabilities, as evidenced by distinct curve separations across renal function categories (all *P*_log-rank_<0.05). Moreover, we discovered that, compared with CVD patients with normal renal function, those with eGFRcr-defined CKD exhibited a 1.1-years earlier onset of PD and a 0.5-years shorter mean survival time. Similarly, those with eGFRcys-defined CKD showed a 0.6-year earlier PD onset and 0.9-years reduction in survival time (Figure S1A-B), respectively.

**Figure 3.**
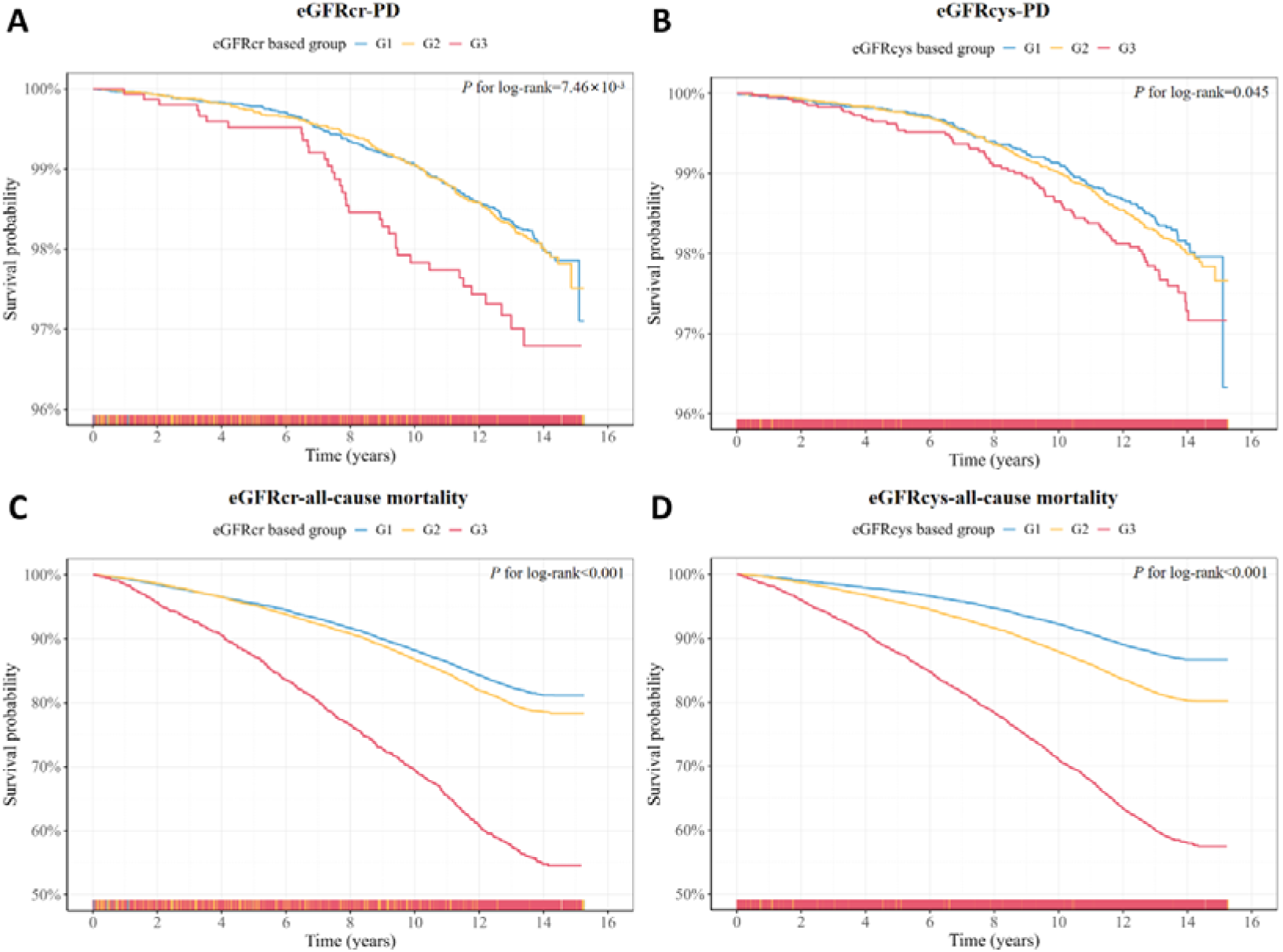
(A) Cumulative incidence of PD among groups of eGFRcr-based renal function categories. (B) Cumulative incidence of PD among groups of eGFRcys-based renal function categories. (C) Cumulative incidence of all-cause mortality among groups of eGFRcr-based renal function categories. (D) Cumulative incidence of all-cause mortality among groups of eGFRcys-based renal function categories. G1, normal renal function group; G2, mildly reduced renal function group; G3, CKD group; PD, Parkinson’s disease; eGFRcr, estimated glomerular filtration rate based on creatinine; eGFRcys, estimated glomerular filtration rate based on cystatin C; CKD, chronic kidney disease.

These patterns were formally validated through Cox proportional hazards regression (Table 2), revealing significant renal risk gradients. Specifically, for incident PD development, eGFRcr-defined CKD was associated with a 60.1% (12.0∼128.9%) elevated hazard, while the corresponding risk increases for eGFRcys-defined CKD was 38.4% (3.0∼85.9%). Notably, mild renal impairment demonstrated no significant association with PD risk for neither eGFRcr- nor eGFRcys-defined categories (both *P*>0.05). Regarding all-cause mortality, eGFRcr-defined mild impairment and CKD respectively heightened death risk by 21.9% (15.4∼28.8%) and 158.9% (137.7∼182.0%). Meanwhile, eGFRcys stratification indicated even more evident associations: mild impairment increased mortality by 50.0% (39.6∼61.2%) and CKD by 233.5% (207.6∼261.7%).

### Sensitivity analyses of renal function-PD link

Sensitivity analyses confirmed consistent relationships between baseline eGFR and risks of incident PD and all-cause mortality (Figure S2 and Table S5). First, a 2-year lag exclusion analysis showed minimal changes in HRs for continuous eGFR and renal function categories, indicating limited potential for reverse causation. Second, the combined GPS/IPW/E-value approach robustly validated the associations against substantial unmeasured confounding. Specifically, GPS modeling balanced baseline confounders across eGFR exposure levels through propensity weighting, while IPW refinement significantly enhanced the precision of mortality estimates, which was most critically for eGFRcys-CKD (HR=3.97 [3.67∼4.30]). Complementarily, all significant E-values consistently exceeded the corresponding HRs (or their reciprocals for HR<1), confirming resilience against residual confounding factors. Third, cause-specific hazards regression confirmed that lower eGFR remained associated with higher PD risk after accounting for competing mortality. Mildly impaired renal function still showed no association with PD risk but maintained substantially stable links with mortality across all these sensitivity analyses.

### Temporal trends of renal function in incident PD and all-cause mortality

The longitudinal trajectories of both eGFR metrics across the 14-year pre-diagnosis/pre-death window are illustrated in Figure 4A-D. In the PD group, both eGFR metrics displayed progressive divergence from matched controls beginning 14 years pre-diagnosis; but a deceleration in decline was noted at years -10 to -5, followed by reversion to control level. In contrast, the all-cause mortality group exhibited consistently lower eGFR levels than the control group, particularly for eGFRcys, which demonstrated a progressive decline from the start of follow-up, with worsening accelerating steeply as the time of death approached. The trajectory-resolved dynamics substantiated time-stratified associations, jointly indicating that subclinical eGFR decline initiates >14 years preclinically for PD and mortality.

**Figure 4.**
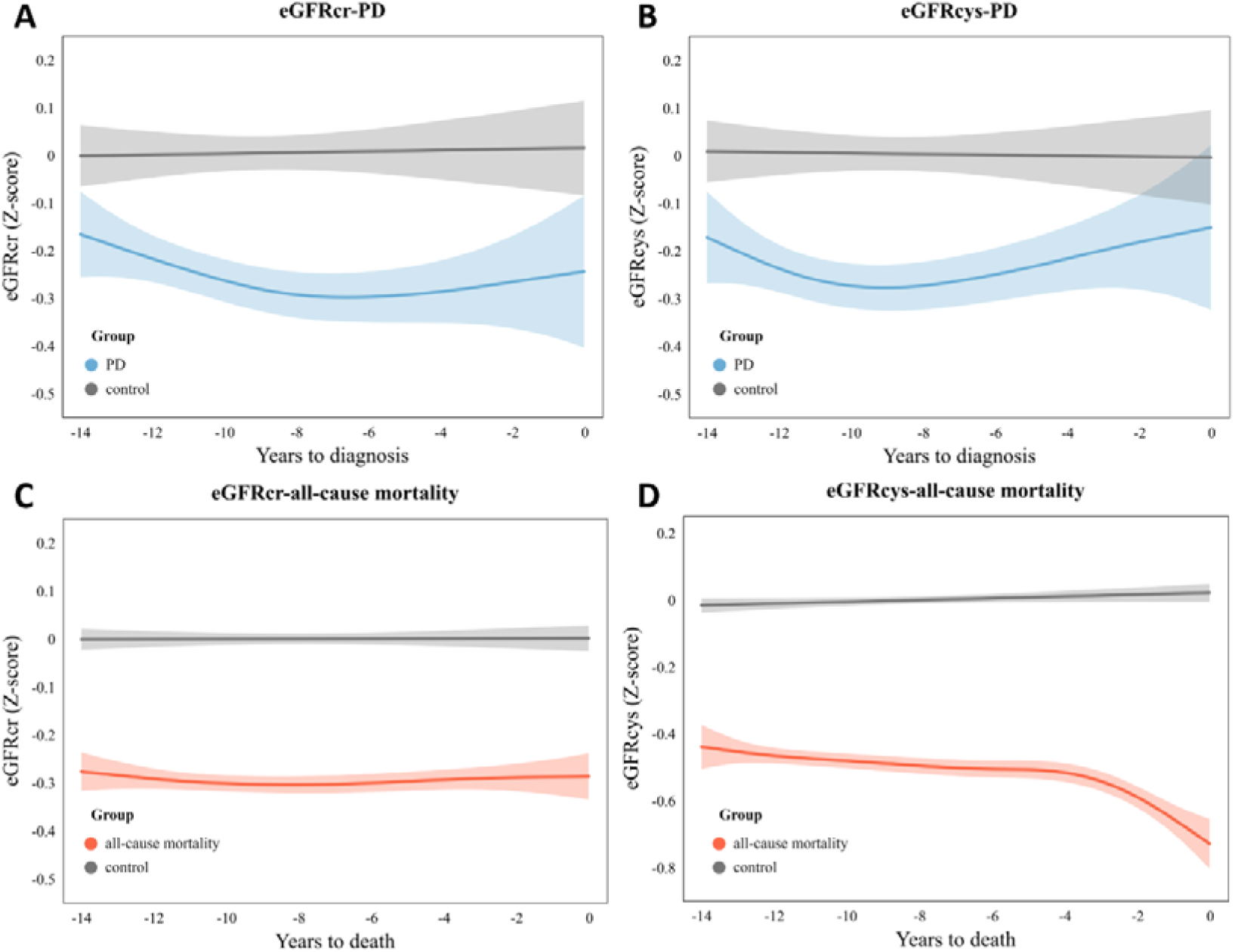
(A) Local weighted scatterplot smoothing of temporal eGFRcr trends before PD diagnosis and control group. (B) Local weighted scatterplot smoothing of temporal eGFRcys trends before PD diagnosis and control group. (C) Local weighted scatterplot smoothing of temporal eGFRcr trends before all-cause death and control group. (D) Local weighted scatterplot smoothing of temporal eGFRcys trends before all-cause death and control group. PD, Parkinson’s disease; eGFRcr, estimated glomerular filtration rate based on creatinine; eGFRcys, estimated glomerular filtration rate based on cystatin C; eGFR, estimated glomerular filtration rate.

### Improvement of renal function on the predictive performance of PREDCIT-PD

The PREDICT-PD algorithm exhibited robust, independent predictive power for PD. Among the three predictive methods evaluated, logistic regression (AUC=0.745 [0.743∼0.747]) outperformed both XGBoost (AUC=0.740 [0.738∼0.742]) and LightGBM (AUC=0.736 [0.734∼0.739]), achieving the highest predictive performance (Figure S2). Consequently, we selected logistic regression for subsequent risk prediction modeling and investigated whether incorporating CKD into PREDICT-PD would improve its predictive capability. After adjustment for *P*_0_, CKD remained a strong, independent predictor of incident PD risk (Table 3; *P*<0.05 across all iterations), with RR_CKD_=1.54 (1.50∼1.58) for eGFRcr-defined CKD and RR_CKD_=1.59 (1.57∼1.62) for eGFRcys-defined CKD. Using RR_CKD_, we derived *P*_1_ and evaluated its predictive ability. Consequently, this new probability achieved notable improvements in discrimination compared with the original one. Specifically, for eGFRcr-defined CKD (AUC=0.754 [0.752∼0.756]), *P*_1_ yielded an absolute increase in AUC of 1.18% (1.16∼1.20%), while for eGFRcys-defined CKD (AUC=0.755 [0.752∼0.757]), the increase was 1.34% (1.32∼1.37%). The continuous NRIs (0.147 [0.137∼0.162] and 0.160 [0.142∼0.186]) and IDIs (0.005 [0.004∼0.006] and 0.006 [0.004∼0.009]) further confirmed the superior incremental discrimination provided by incorporating renal function into the PD risk prediction algorithm.

**Table 3.**
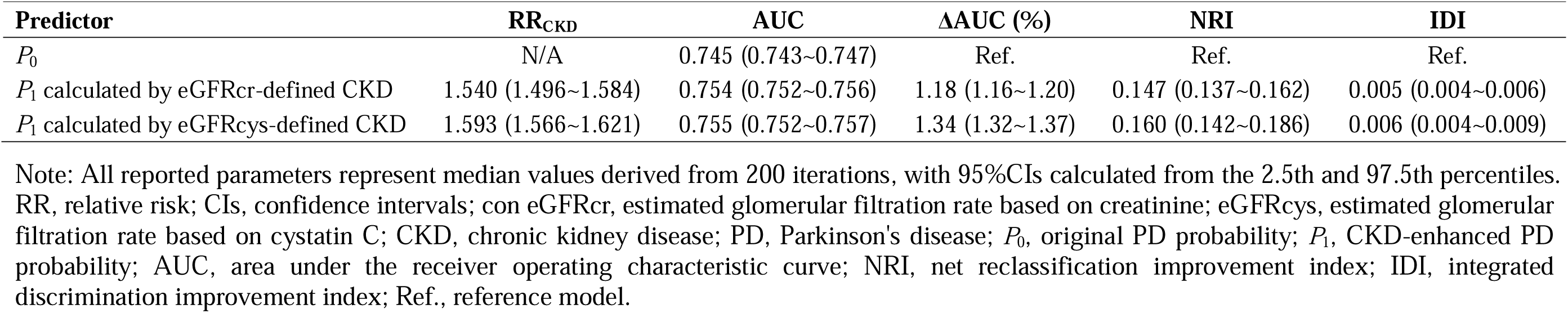
Predictive capacity of original and CKD enhanced model of PREDICT-PD on PD.

| Predictor | RR<sub>CKD</sub> | AUC | ΔAUC (%) | NRI | IDI |
| --- | --- | --- | --- | --- | --- |
| *P*<sub>0</sub> | N/A | 0.745 (0.743~0.747) | Ref. | Ref. | Ref. |
| *P*<sub>1</sub> calculated by eGFR<sub>cr</sub>-defined CKD | 1.540 (1.496~1.584) | 0.754 (0.752~0.756) | 1.18 (1.16~1.20) | 0.147 (0.137~0.162) | 0.005 (0.004~0.006) |
| *P*<sub>1</sub> calculated by eGFR<sub>cys</sub>-defined CKD | 1.593 (1.566~1.621) | 0.755 (0.752~0.757) | 1.34 (1.32~1.37) | 0.160 (0.142~0.186) | 0.006 (0.004~0.009) |

Note: All reported parameters represent median values derived from 200 iterations, with 95% CIs calculated from the 2.5th and 97.5th percentiles. RR, relative risk; CIs, confidence intervals; con eGFR<sub>cr</sub>, estimated glomerular filtration rate based on creatinine; eGFR<sub>cys</sub>, estimated glomerular filtration rate based on cystatin C; CKD, chronic kidney disease; PD, Parkinson's disease; *P*<sub>0</sub>, original PD probability; *P*<sub>1</sub>, CKD-enhanced PD probability; AUC, area under the receiver operating characteristic curve; NRI, net reclassification improvement index; IDI, integrated discrimination improvement index; Ref., reference model.

## Discussion

### Restatement of the core findings

Our study provided robust evidence linking impaired renal function, measured by both eGFRcr and eGFRcys, to an elevated risk of incident PD and all-cause mortality within a large cohort of individuals with pre-existing CVD. The significant inverse associations observed for both eGFR metrics demonstrate that reduced renal function is an independent risk factor for PD and all-cause mortality in this high-risk population. Further, our longitudinal trajectory analysis revealed a slow, progressive decline in eGFR begins over 14 years prior to PD diagnosis, establishing renal dysfunction as a protracted, prodromal process rather than a mere consequence of established disease. Critically, incorporating eGFR-defined CKD status significantly enhanced the discrimination and prediction accuracy of the conventional PREDICT-PD algorithm in individuals with CVD using only routine clinical data. These findings reveal renal impairment as both a biomarker and a target for mitigating PD and all-cause mortality risk in vulnerable CVD patients.

### Engagement with prior research

#### Enhanced renal-PD association in the CVD-specific population

Our study extends and validates prior findings available from studies conducted in general populations 4–6,32 by demonstrating that declined renal function substantially elevates PD risk specifically within the CVD patient cohort. Crucially, we provide evidence indicating an earlier manifestation of this risk and a pronounced effect compared with the general population. Unlike Peng et al. 6, who only observed increased PD risk primarily in cases of severe renal impairment (eGFR<30 ml/min/1.73m²) in a general population cohort, our findings reveal significant associations in the CVD population at an earlier stage of renal decline (eGFR<60 ml/min/1.73m²). Furthermore, while Kwon et al. documented a relatively modest CKD-associated PD risk increase in the general population (HR=1.20 [1.04∼1.39]) 4, our analysis within the CVD patient cohort demonstrates evidently greater risks, particularly for eGFRcr-defined CKD (HR=1.60 [1.12∼2.29]). These observations support our hypothesis that dysregulated cardiorenal crosstalk in CVD populations exacerbates renal dysfunction’s neuropathological impact, amplifying the detrimental impact of renal dysfunction and culminating in a further elevation of PD incidence.

#### Avoidance of inherent robustness issues in conventional analyses

Observational studies linking renal dysfunction to PD face inherent epistemological challenges: reverse causation can misattribute effects, unmeasured confounders muddy pathophysiology, and competing mortality risk can distort associations in comorbid cohorts 33,34. Our tailored sensitivity framework transcends these limitations to strengthen causal attribution. The preserved hazard ratios after rigorous temporal exclusions affirm renal impairment’s role as a preclinical antecedent, not a neurodegenerative consequence. By applying GPS 13 and IPW 14 for confounder adjustment across continuous renal exposures, we isolated biological signals obscured in conventional models, while quantitative E-value analysis 14 confirmed that unmeasured biases would require substantially strong effects to negate our findings. Critically, explicit competing risk adjustment verifies that PD’s risk persists independently of heightened mortality — addressing a fundamental validity threat in cardio-renal cohorts. Through these multi-angle sensitivity analyses, we robustly validated the links observed.

#### Early renal decline signals higher PD risk decades before onset

Our temporal analysis provides novel insights into the long-term temporal association between renal function decline and PD onset among individuals with CVD. Observational studies often report associations without capturing trajectories; in contrast, by mapping eGFR trajectories over 14 years pre-diagnosis through nested case-control analysis, we show a progressive divergence in renal function starting more than a decade before the clinical recognition of PD. This protracted subclinical decline suggests shared progressive pathophysiological mechanisms operating long before symptomatic disease onset, potentially involving neuroinflammatory pathways 35, uremic toxin accumulation 36,37, or systemic metabolic dysfunction 38,39. This finding both aligns with the known prodromal phase of PD (10∼15 years) 40, indicating renal dysfunction could be an early biomarker within this window for individuals with CVD.

#### Predictive enhancement through renal function refinement

Within CVD populations, a group at elevated risk for PD yet underrepresented in existing prediction models, clinically feasible risk estimation remains challenging. Here, in this high-risk individuals, we demonstrated that leveraging CKD defined by routinely available eGFR significantly enhanced the predictive performance (ΔAUC: 1.18∼1.30%) of PREDICT-PD 17, an established general-population PD risk prediction model. Crucially, after rigorously comparing three distinct modeling approaches (logistic regression, XGBoost, and LightGBM), we identified logistic regression as delivering the most robust predictive value upon the integration of eGFR-defined CKD. Through cross-validated optimization, logistic regression effectively captured the renal dysfunction-PD risk association, leveraging its inherent interpretability and strong linear predictive power — attributes highly aligned with clinical application needs.

This focused enhancement fundamentally contrasts with emerging biomarker strategies. Recent general-population PD risk prediction models (e.g., Danek et al.’s multi-omics model 41) demand costly, specialized assays unavailable in routine practice. To achieve comparable performance specifically in the CVD subpopulation, such methods would require further validation and potential recalibration in CVD cohorts, and persistent reliance on expensive, non-routine testing. Thus, by repurposing ubiquitous clinical data uniquely accessible within CVD management contexts, our CKD-enhanced model provides a rapidly implementable, highly cost-effective advance tailored to real-world cardiovascular care constraints 21.

#### Renal impairment behind the high mortality in CVD patients

The profound association between impaired renal function, particularly CKD defined by eGFRcys, and all-cause mortality within our CVD cohort warrants specific emphasis. The substantially larger hazard ratios observed for mortality compared with PD (e.g., eGFRcys-defined CKD: HR=3.34 for all-cause mortality vs. 1.38 for PD) underscore that renal health is a dominant determinant of survival in this high-risk population. This association aligns with and significantly extends the well-established cardiorenal paradigm 42, demonstrating that severe renal dysfunction portends excess all-cause mortality risk independent of underlying cardiovascular pathology 43. The robustness of this association, confirmed by multiple sensitivity analyses, highlights routine assessment of renal function, especially eGFRcys where feasible, as essential for comprehensive prognosis in CVD management.

Our longitudinal trajectory analysis provides novel mechanistic context: eGFR decline associated with mortality was progressive and accelerated steeply in the years preceding death, contrasting with the relatively slower and plateauing pre-PD trends. This acceleration phase, especially evident using eGFRcys suggests a confluence of systemic deterioration and escalating inflammatory/hemodynamic stress near the end of life 44,45. The shared pathways implicated in both renal-mortality (accelerated cardiorenal decline) and renal-PD (chronic neurotoxicity/inflammation) links, namely uremic toxin accumulation, chronic inflammation, oxidative stress, and dysautonomia 7,36,37, further support the central role of aberrant cardiorenal crosstalk in driving multifaceted adverse outcomes in this vulnerable group.

### Prevention, prediction and monitoring

Our findings have significant public health and clinical implications for managing patients with CVD, a population already at high risk. Incorporating readily available CKD status into the established PREDICT-PD framework 17 demonstrably enhances PD risk prediction, offering clinicians a practical tool. Identifying CVD patients with concomitant CKD flags them for increased surveillance and potentially earlier neurological evaluation should prodromal PD symptoms emerge 1. Crucially, this subgroup also faces >3-fold higher all-cause mortality risk associated with eGFRcys-defined CKD. Thus, monitoring renal function serves dual purposes: as an early sentinel for developing PD pathology decades before motor symptoms, and as a critical survival indicator in this high-risk cohort.

While eGFR alone lacks diagnostic specificity for PD, its sustained unexplained decline in CVD patients warrants vigilance for both neurological and systemic complications 46. Public health strategies prioritizing renal health within CVD management may thus concurrently mitigate PD risk and reduce premature mortality, especially where eGFRcys assessment is feasible. This association provides crucial insight into the complex comorbidity profile of CVD patients, underscoring the need to consider renal health within the broader context of systemic degeneration and mortality risk. Further research should explore if optimizing renal health modifies PD risk, progression, or survival in susceptible individuals.

### Limitations of our research

Our findings must be interpreted within the context of the study’s limitations. First, the restriction to individuals of European ancestry ensures population homogeneity but limits generalizability to other ethnic groups known to have different CKD prevalences and PD risks 47. Second, serum creatinine and cystatin C levels, while widely used to estimate GFR, do not completely equate to measured GFR 21. Although we have comprehensively examined their associations with PD risk and mortality at the phenotypic level, causal evidence remains limited; further mechanistic investigations and intervention trials are warranted to establish causality 48,49. Third, our prediction enhancement was implemented within the specific PREDICT-PD algorithm 17; the magnitude of improvement might differ if applied to other prediction models. Finally, this study focused on prevalent CVD patients, so results may not directly apply to healthy populations or those without CVD.

### Conclusions

This work provides compelling evidence that impaired renal function, particularly CKD, is a significant independent risk factor for incident PD and all-cause mortality among individuals with pre-existing CVD. This association manifests as a slow, progressive decline in renal function initiating more than a decade prior to PD diagnosis and is not substantially affected by confounding, reverse causality, or competing mortality risks. The practical utility of renal function is underscored by its ability to significantly enhance the performance of the established PREDICT-PD prediction algorithm. These findings highlight renal dysfunction as an integral component of the systemic prodromal phase of PD, with profound implications for both understanding pathophysiological mechanisms linking the renal and the brain and improving early risk stratification strategies in high-risk subpopulations.

## Data Availability

Data sharing not applicable to this article as no datasets were generated or analyzed during the current study.

## List of abbreviations

PD: Parkinson’s disease
CKD: Chronic kidney disease
eGFR: Estimated glomerular filtration rate
CVD: Cardiovascular disease
eGFRcr: Creatinine-based estimated glomerular filtration rate
eGFRcys: Cystatin C-based estimated glomerular filtration rate
PSM: Propensity score matching
EHR: Electronic health record
TDI: Thompson deprivation index
BMI: Body mass index
SD: Standard deviation
HR: Hazard ratio
CI: Confidence interval
GPS: Generalized propensity score
IPW: Inverse probability weight
RR: Relative risk
AUC: Area under the receiver operating characteristic curve
NRI: Net reclassification improvement index
IDI: Integrated discrimination improvement index

## Additional File

Supplementary File.

## Ethics approval and consent to participate

Not applicable.

## Consent for publication

Not applicable.

## Authors’ contributions

P.Z. conceived the idea for the study. P.Z. obtained the data. P.Z. and J.Q. cleared up the datasets; P.Z. and J.Q. performed the data analyses. P.Z., and J.Q. interpreted the results of the data analyses. P.Z. and J.Q. wrote the manuscript with the help from other authors.

## Acknowledgements

This study was mainly based on the UK Biobank resource under application number 88159. The UK Biobank was established by the Wellcome Trust medical charity, Medical Research Council, Department of Health, Scottish Government, and the Northwest Regional Development Agency. It has also had funding from the Welsh Assembly Government, British Heart Foundation and Diabetes UK. The data analyses in the present study were carried out with the high-performance computing cluster that was supported by the special central finance project of local universities for Xuzhou Medical University.

