## Supplementary Files for "Renal Impairment and Parkinson’s Disease in Cardiovascular Patients: Associations, Pre-diagnostic Trajectories, and Predictive Enhancement"

**Supplementary File**

**Treatment of exposures and outcomes**

We defined cardiovascular disease (CVD) and Parkinson's disease (PD) by ICD-10. We then used serum creatinine concentration, serum cystatin C concentration, gender and age at recruitment (field 21022, continuous) to calculate two types of estimated glomerular filtration rate (eGFR; eGFRcr: eGFR based on serum creatinine; eGFRcys: eGFR based on cystatin C) in terms of the currently recommended CKD-EPI equation [1, 2]. The creatinine-based eGFR (eGFRcr, unit: ml/min/1.73m^2^) was computed using the formula:

$$\text{eGFRcr=142×}\left( \frac{\text{Scr}}{\text{A}} \right)\text{B}\text{×0.9938}\text{age}\text{×(1.012 if female)}$$

where Scr represents serum creatinine concentration (unit: mg/dL). For female participants, parameter assignments were as follows: when Scr ≤0.7 mg/dL, *A* was set to 0.7 and *B* to -0.241; if Scr >0.7 mg/dL, *A* remained 0.7 while *B* was adjusted to -1.2. For male participants, thresholds differed: when Scr ≤0.9 mg/dL, *A* was 0.9 and *B* was -0.302; for Scr >0.9 mg/dL, *A* remained 0.9, and *B* was changed to -1.2. The cystatin C-based eGFR (eGFRcys, unit: ml/min/1.73m^2^) was calculated as:

$$\text{eGFRcys=133×}\left( \frac{\text{Scys}}{\text{0.8}} \right)\text{A}\text{×0.996}\text{age}\text{×}\text{B}$$

where Scys denotes serum cystatin C concentration (unit: mg/L). For female participants, parameters were assigned as follows: when Scys ≤0.8 mg/L, *A* was -0.499 and *B* was 0.932; if Scys >0.8 mg/L, *A* was adjusted to -1.328 while *B* remained 0.932. For male participants, when Scys ≤0.8 mg/L, *A* was -0.499 and *B* was 1; when Scys >0.8 mg/L, *A* became -1.328 and *B* remained 1. Lower eGFR values reflect a decline in renal function.

**Construction of PREDICT-PD algorithm**

The PREDICT-PD algorithm [3] calculates baseline PD odds (*r*_0_) based on age:

$$\text{r}\text{0}\text{=}\frac{1}{\text{28.53049+73.67057}e^{\text{-0.165308}\left( \text{age-60} \right)}}$$

This *r*_0_ is then multiplicatively adjusted by factors derived from published meta-analyses: divided by 1.5 for female, multiplied by 0.44 for current smoking, 0.78 for previous smoking, 4.45 for positive family history, 0.67 for drinking >1 cup of coffee/day, 0.90 for drinking >1 alcoholic beverage/week, 0.74 for hypertension, 0.83 for non-steroidal anti-inflammatory drugs use, 0.90 for calcium channel blockers use, 1.28 for *β*-blocker use, 2.34 for constipation, 1.58 for head injury, 1.86 for anxiety or depression, and 3.80 for erectile dysfunction.

Table S1. Definitions for all related variables.

| **Variable** | **UK Biobank field ID** | **ICD-10 Code** |
| --- | --- | --- |
| Cardiovascular disease | 41270 | I20-I25, I60-I64, I69 |
| Serum creatinine concentration | 30700 | N/A |
| Serum cystatin C concentration | 30720 | N/A |
| Parkinson's disease (PD) | 41270 | G20 |
| Type 2 diabetes | 41270 | E11 |
| Hypertension | 41270 | I10 |
| Dyslipidemia | 41270 | E78 |
| Gender | 31 | N/A |
| Age at recruitment | 21022 | N/A |
| Body mass index | 21001 | N/A |
| Household income | 738 | N/A |
| Townsend deprivation index | 189 | N/A |
| Education level | 6138 | N/A |
| Smoking status | 20116 | N/A |
| Alcohol consumption | 1558 | N/A |
| Physical activity level | 22032 | N/A |
| Healthy diet score | 1289, 1299, 1309, 1319, 1329, 1339, 1349, 1369, 1379, 1389 | N/A |
| Medication use | 20003 | N/A |
| Top 10 genetic principal components | 22009 | N/A |
| Family history of PD | 20107, 20110, 20111 | N/A |
| Coffee consumption | 1498 | N/A |
| Constipation | 41270 | K59.0 |
| Head injury | 41270 | S02.0, S02.1, S02.2, S02.3, S02.4, S02.6, S02.7, S02.8, S02.9, S06.0, S06.1, S06.2, S06.3, S06.4, S06.5, S06.6, S06.8, S06.9 |
| Anxiety | 41270 | F41.2, F41.3, F41.8, F41.9 |
| Depression | 41270 | F32.0, F32.1, F32.2, F32.3, F32.8, F32.9, F33.0, F33.1, F33.2, F33.3, F33.4, F33.8, F33.9 |
| Erectile dysfunction | 41270 | N48.4 |

Table S2. Definitions of top-frequency CVD medication use.

| **Code** | **Drug name** |
| --- | --- |
| **ACEIs** |  |
| 1140860750 | captopril |
| 1140860764 | captopril+hydrochlorothiazide 25mg/12.5mg tablet |
| 1140860758 | capoten 12.5mg tablet |
| 1141150328 | ecopace 12.5mg tablet |
| 1141167758 | hyteneze 12.5 tablet |
| 1141150560 | kaplon 12.5mg tablet |
| 1141151382 | hypapril 12.5mg tablet |
| 1140881714 | capozide tablet |
| 1140888552 | enalapril |
| 1140866340 | delvas tablet |
| 1140860790 | enalapril maleate+hydrochlorothiazide 20mg/12.5mg tablet |
| 1140860776 | innovace 2.5mg tablet |
| 1141170870 | pralenal 2.5mg tablet |
| 1140881712 | renitec 5mg tablet |
| 1140860784 | innozide tablet |
| 1140864952 | lisinopril+hydrochlorothiazide 10mg/12.5mg tablet |
| 1140860752 | acepril 12.5mg tablet |
| 1140860696 | lisinopril |
| 1140860714 | zestril 2.5mg tablet |
| 1140864618 | zestoretic 10 tablet |
| 1140888560 | perindopril |
| 1141180592 | perindopril+indapamide |
| 1140860802 | coversyl 2mg tablet |
| 1141180598 | coversyl plus 4mg/1.25mg tablet |
| 1140860806 | ramipril |
| 1141165470 | felodipine+ramipril |
| 1141188408 | tritace 1.25mg tablet |
| 1141199940 | lopace 2.5mg capsule |
| 1141200698 | ranace 1.25mg capsule |
| 1140860706 | carace 2.5mg tablet |
| 1140864910 | carace 10 plus tablet |
| 1141165476 | triapin mite 2.5mg/2.5mg tablet |
| 1140860738 | quinalapril+hydrochlorothiazide 10mg/12.5mg tablet |
| 1140860728 | quinapril |
| 1140881706 | accupro 5mg tablet |
| 1140860736 | accuretic tablet |
| 1140860882 | cilazapril |
| 1140860892 | vascace 250micrograms tablet |
| 1140888556 | fosinopril |
| 1140864176 | monozide 10 tablet |
| 1140860878 | staril 10mg tablet |
| 1140860904 | trandolapril |
| 1141153328 | trandolapril + verapamil hydrochloride |
| 1140860912 | gopten 500micrograms capsule |
| 1140860918 | odrik 500micrograms capsule |
| 1141153316 | tarka 2mg/180mg m/r capsule |
| 1140923712 | moexipril |
| 1140923718 | perdix 7.5mg tablet |
| 1141164148 | imidapril hydrochloride |
| 1141164154 | tanatril 5mg tablet |
| **ARBs** |  |
| 1140916356 | losartan |
| 1141151016 | losartan potassium+hydrochlorothiazide 50mg/12.5mg tablet |
| 1140916362 | cozaar half strength 25mg tablet |
| 1141151018 | cozaar-comp 50mg/12.5mg tablet |
| 1141179974 | cozaar 25mg tablet |
| 1141171336 | eprosartan |
| 1141171344 | teveten 300mg tablet |
| 1141145660 | valsartan |
| 1141201038 | valsartan+hydrochlorothiazide 80mg/12.5mg tablet |
| 1141145668 | diovan 40mg capsule |
| 1141201040 | co-diovan 80mg/12.5mg tablet |
| 1141172682 | irbesartan+hydrochlorothiazide 150mg/12.5mg tablet |
| 1141152998 | irbesartan |
| 1141153006 | aprovel 75mg tablet |
| 1141172686 | coaprovel 150mg/12.5mg tablet |
| 1141156836 | candesartan cilexetil |
| 1141156846 | amias 2mg tablet |
| 1141166006 | telmisartan |
| 1141187788 | telmisartan+hydrochlorothiazide 40mg/12.5mg tablet |
| 1141187790 | micardisplus 40mg/12.5mg tablet |
| 1141172492 | micardis 20mg tablet |
| 1141193282 | olmesartan |
| 1141193346 | olmetec 10mg tablet |
| **BBs** |  |
| 1140860212 | apsolox 20mg tablet |
| 1140851576 | laracor 20mg tablet |
| 1140879830 | oxprenolol |
| 1140860334 | trasidrex tablet |
| 1140860230 | oxyprenix sr 160mg m/r tablet |
| 1140851484 | paritane 20mg tablet |
| 1140851480 | slow-pren 160mg m/r tablet |
| 1140860220 | slow-trasicor 160mg m/r tablet |
| 1140860222 | trasicor 20mg tablet |
| 1140860292 | pindolol |
| 1140860322 | pindolol+clopamide 10mg/5mg tablet |
| 1140910614 | prindolol |
| 1140860338 | viskaldix tablet |
| 1140860294 | visken 5mg tablet |
| 1140866704 | angilol 10mg tablet |
| 1140866764 | apsolol 10mg tablet |
| 1140851556 | bedranol 10mg tablet |
| 1140866784 | berkolol 10mg tablet |
| 1140866782 | beta-prograne 160mg m/r capsule |
| 1140866778 | betadur cr 160mg m/r capsule |
| 1140866712 | cardinol 10mg tablet |
| 1140866802 | half beta-prograne 80mg m/r capsule |
| 1141152076 | half propanix la 80mg m/r capsule |
| 1141156754 | half propatard la 80mg m/r capsule |
| 1140866798 | half-betadur cr 80mg m/r capsule |
| 1140866800 | half-inderal la 80mg m/r capsule |
| 1140866804 | inderal 10mg tablet |
| 1140917076 | lopranol la 160mg m/r capsule |
| 1140866766 | propanix 10mg tablet |
| 1141156808 | propatard la 160mg m/r capsule |
| 1140879842 | propranolol |
| 1140860418 | propranolol hydrochloride+bendrofluazide 80mg/2.5mg capsule |
| 1141187048 | rapranol sr 80mg m/r capsule |
| 1140916730 | sloprolol 80mg m/r capsule |
| 1141172742 | syprol 5mg/5ml oral solution |
| 1140881722 | avlocardyl retard 160mg m/r capsule |
| 1140851508 | spiroprop tablet |
| 1140879866 | timolol |
| 1140860340 | timolol maleate+bendrofluazide 10mg/2.5mg tablet |
| 1140860342 | timolol maleate+bendrofluazide 20mg/5mg tablet |
| 1141194808 | timolol maleate+bendroflumethiazide 10mg/2.5mg tablet |
| 1140860336 | timolol maleate+co-amilozide 10mg/2.5mg/25mg tablet |
| 1140879854 | sotalol |
| 1140860304 | beta-cardone 40mg tablet |
| 1140860362 | sotacor 80mg tablet |
| 1140860332 | sotalol hydrochloride+hydrochlorothiazide 80mg/12.5mg tablet |
| 1140860318 | sotazide tablet |
| 1140860194 | corgard 40mg tablet |
| 1140860192 | nadolol |
| 1140860312 | nadolol+bendrofluazide 40mg/5mg tablet |
| 1140860316 | nadolol+bendrofluazide 80mg/5mg tablet |
| 1141194804 | nadolol+bendroflumethiazide 40mg/5mg tablet |
| 1140879822 | carteolol |
| 1140879834 | penbutolol |
| 1140860320 | penbutolol sulphate+furosemide 40mg/20mg tablet |
| 1140860180 | arbralene 50mg tablet |
| 1140860266 | betaloc 50mg tablet |
| 1140860386 | co-betaloc tablet |
| 1140860274 | lopresor 50mg tablet |
| 1140860402 | lopresoretic tablet |
| 1140860278 | mepranix 50mg tablet |
| 1140879818 | metoprolol |
| 1140860308 | metoprolol tartrate+chlorthalidone 100mg/12.5mg tablet |
| 1140860404 | metoprolol tartrate+hydrochlorothiazide 100mg/12.5mg tablet |
| 1140851522 | metoros 95mg tablet |
| 1141182968 | tensomex 100mg tablet |
| 1140866738 | atenolol |
| 1141146126 | atenolol+bendrofluazide |
| 1141194810 | atenolol+bendroflumethiazide |
| 1141180778 | atenolol+chlortalidone |
| 1141146124 | atenolol+chlorthalidone |
| 1141146128 | atenolol+co-amilozide |
| 1140922930 | atenix 25mg tablet |
| 1140860348 | atenixco 50mg/12.5mg tablet |
| 1140864410 | antipressan 25mg tablet |
| 1140860172 | totamol 25mg tablet |
| 1140866758 | vasaten 50mg tablet |
| 1140923336 | co-tenidone |
| 1140860324 | tenoret 50 tablet |
| 1140860328 | tenoretic tablet |
| 1140866756 | tenormin 25 tablet |
| 1140860426 | atenolol+nifedipine 50mg/20mg m/r capsule |
| 1140860358 | tenif capsule |
| 1140860356 | beta-adalat capsule |
| 1140860398 | kalten capsule |
| 1140866724 | acebutolol |
| 1140860422 | acebutolol+hydrochlorothiazide 200mg/12.5mg tablet |
| 1140866726 | sectral 100mg capsule |
| 1140879758 | betaxolol |
| 1140851492 | betadren 5mg tablet |
| 1141184324 | bipranix 5mg tablet |
| 1140879760 | bisoprolol |
| 1140864950 | bisoprolol fumarate+hydrochlorothiazide 10mg/6.25mg tablet |
| 1141171152 | cardicor 1.25mg tablet |
| 1140860492 | emcor 10mg tablet |
| 1140860434 | monocor 5mg tablet |
| 1141182904 | soloc 5mg tablet |
| 1141187780 | vivacor 5mg tablet |
| 1140879762 | celiprolol |
| 1140860498 | celectol 200mg tablet |
| 1141164280 | nebilet 5mg tablet |
| 1141164276 | nebivolol |
| 1140879824 | labetalol |
| 1140860244 | labrocol 100mg tablet |
| 1140860250 | trandate 50mg tablet |
| 1140909368 | carvedilol |
| 1141168498 | eucardic 3.125 tablet |
| **CCBs** |  |
| 1140879802 | amlodipine |
| 1141200400 | amlostin 5mg tablet |
| 1140861202 | istin 5mg tablet |
| 1141187094 | cabren 2.5mg m/r tablet |
| 1141199858 | cardioplen xl 5mg m/r tablet |
| 1141188836 | felendil xl 5mg m/r tablet |
| 1140888646 | felodipine |
| 1141165470 | felodipine+ramipril |
| 1141188576 | felogen xl 5mg m/r tablet |
| 1141188152 | felotens xl 5mg m/r tablet |
| 1141188920 | keloc sr 5mg m/r tablet |
| 1141200782 | neofel xl 5mg m/r tablet |
| 1140868036 | parmid 10mg tablet |
| 1141201814 | parmid xl 5mg m/r tablet |
| 1140928212 | plendil 2.5mg m/r tablet |
| 1141190160 | vascalpha 5mg m/r tablet |
| 1141165476 | triapin mite 2.5mg/2.5mg tablet |
| 1140861190 | isradipine |
| 1140861194 | prescal 2.5mg tablet |
| 1140861176 | cardene 20mg capsule |
| 1140879810 | nicardipine |
| 1140860426 | atenolol+nifedipine 50mg/20mg m/r capsule |
| 1140860358 | tenif capsule |
| 1140861090 | adalat 5mg capsule |
| 1140881702 | adalate 10mg capsule |
| 1140923572 | adipine mr 10 m/r tablet |
| 1140861110 | angiopine 5mg capsule |
| 1140860356 | beta-adalat capsule |
| 1140916930 | calanif 5mg capsule |
| 1141173766 | calchan mr 10mg m/r tablet |
| 1140861106 | calcilat 10mg capsule |
| 1140927934 | cardilate mr 10mg m/r tablet |
| 1140861120 | coracten sr 10mg m/r capsule |
| 1141166752 | coroday mr 20mg m/r tablet |
| 1141145870 | fortipine la40 m/r tablet |
| 1141152600 | genalat retard 10mg m/r tablet |
| 1141187962 | kentipine mr 10mg m/r tablet |
| 1140861088 | nifedipine |
| 1141157140 | nifedipress mr 10 m/r tablet |
| 1141150538 | nifedotard 20mr m/r tablet |
| 1140911088 | nifelease 20mg m/r tablet |
| 1140861114 | nifensar xl 20mg m/r tablet |
| 1141169730 | nifopress retard 20mg m/r tablet |
| 1140926966 | nimodrel mr 10 m/r tablet |
| 1141162546 | nivaten retard 10mg m/r tablet |
| 1141150500 | slofedipine 20mg m/r tablet |
| 1140927940 | tensipine mr 10 m/r tablet |
| 1140926188 | unipine xl 30mg m/r tablet |
| 1141190548 | valni 20 retard 20mg m/r tablet |
| 1140851790 | vasad 5mg capsule |
| 1140872568 | nimodipine |
| 1140872472 | nimotop 30mg tablet |
| 1140928226 | nisoldipine |
| 1140928234 | syscor mr 10mg m/r tablet |
| 1140861276 | lacidipine |
| 1140861282 | motens 2mg tablet |
| 1141153026 | lercanidipine |
| 1141153032 | zanidip 10mg tablet |
| **Diuretics** |  |
| 1140866324 | triamterene+benzthiazide 50mg/25mg capsule |
| 1141194794 | bendroflumethiazide |
| 1141194800 | bendroflumethiazide+potassium 2.5mg/7.7mmol m/r tablet |
| 1140866440 | centyl k m/r tablet |
| 1140851332 | centyl 2.5mg tablet |
| 1141194804 | nadolol+bendroflumethiazide 40mg/5mg tablet |
| 1141194808 | timolol maleate+bendroflumethiazide 10mg/2.5mg tablet |
| 1141194810 | atenolol+bendroflumethiazide |
| 1140866136 | neo-naclex 5mg tablet |
| 1140866446 | neo-naclex k m/r tablet |
| 1140866128 | aprinox 2.5mg tablet |
| 1140866122 | bendrofluazide |
| 1140860312 | nadolol+bendrofluazide 40mg/5mg tablet |
| 1140860316 | nadolol+bendrofluazide 80mg/5mg tablet |
| 1140860340 | timolol maleate+bendrofluazide 10mg/2.5mg tablet |
| 1140860342 | timolol maleate+bendrofluazide 20mg/5mg tablet |
| 1140860418 | propranolol hydrochloride+bendrofluazide 80mg/2.5mg capsule |
| 1140866450 | bendrofluazide+potassium 2.5mg/7.7mmol m/r tablet |
| 1140910442 | bzt - bendrofluazide |
| 1141146126 | atenolol+bendrofluazide |
| 1140866132 | berkozide 2.5mg tablet |
| 1140888918 | neo-bendromax 2.5mg tablet |
| 1140851336 | urizide 5mg tablet |
| 1140923282 | co-flumactone |
| 1140866396 | aldactide 25 tablet |
| 1140866354 | amilmaxco 5/50 tablet |
| 1141187790 | micardisplus 40mg/12.5mg tablet |
| 1141151018 | cozaar-comp 50mg/12.5mg tablet |
| 1141172686 | coaprovel 150mg/12.5mg tablet |
| 1141201040 | co-diovan 80mg/12.5mg tablet |
| 1140851430 | synuretic tablet |
| 1140851432 | hypertane-50 tablet |
| 1140866360 | triamaxco tablet |
| 1140866328 | triam-co tablet |
| 1140860784 | innozide tablet |
| 1140860736 | accuretic tablet |
| 1140866162 | hydrochlorothiazide |
| 1140860332 | sotalol hydrochloride+hydrochlorothiazide 80mg/12.5mg tablet |
| 1140926778 | diltiazem hcl+hydrochlorothiazide 150mg/12.5mg m/r capsule |
| 1141151016 | losartan potassium+hydrochlorothiazide 50mg/12.5mg tablet |
| 1140860404 | metoprolol tartrate+hydrochlorothiazide 100mg/12.5mg tablet |
| 1140860422 | acebutolol+hydrochlorothiazide 200mg/12.5mg tablet |
| 1140860386 | co-betaloc tablet |
| 1140860562 | methyldopa+hydrochlorothiazide 250mg/15mg tablet |
| 1140860738 | quinalapril+hydrochlorothiazide 10mg/12.5mg tablet |
| 1140860764 | captopril+hydrochlorothiazide 25mg/12.5mg tablet |
| 1140860790 | enalapril maleate+hydrochlorothiazide 20mg/12.5mg tablet |
| 1140864950 | bisoprolol fumarate+hydrochlorothiazide 10mg/6.25mg tablet |
| 1140864952 | lisinopril+hydrochlorothiazide 10mg/12.5mg tablet |
| 1141172682 | irbesartan+hydrochlorothiazide 150mg/12.5mg tablet |
| 1141187788 | telmisartan+hydrochlorothiazide 40mg/12.5mg tablet |
| 1141201038 | valsartan+hydrochlorothiazide 80mg/12.5mg tablet |
| 1140851362 | esidrex k tablet |
| 1140851660 | serpasil-esidrex tablet |
| 1140866164 | esidrex 25mg tablet |
| 1140866168 | hydrosaluric 25mg tablet |
| 1140864176 | monozide 10 tablet |
| 1140860318 | sotazide tablet |
| 1140923276 | co-amilozide |
| 1141146128 | atenolol+co-amilozide |
| 1140851436 | vasetic co-amilozide 5/50mg tablet |
| 1140860336 | timolol maleate+co-amilozide 10mg/2.5mg/25mg tablet |
| 1140866420 | moduretic tablet |
| 1140866416 | moduret 25 tablet |
| 1140866402 | dyazide tablet |
| 1140923272 | co-triamterzide |
| 1140860398 | kalten capsule |
| 1140881714 | capozide tablet |
| 1140864618 | zestoretic 10 tablet |
| 1140866138 | chlorothiazide |
| 1140866102 | polythiazide |
| 1140866104 | nephril 1mg tablet |
| 1140866156 | cyclopenthiazide |
| 1140866352 | navispare tablet |
| 1140851368 | navidrex-k tablet |
| 1140866158 | navidrex 500mcg tablet |
| 1140866422 | amiloride hcl+cyclopenthiazide 2.5mg/250micrograms tablet |
| 1140860334 | trasidrex tablet |
| 1140866090 | methyclothiazide |
| 1140851338 | enduron 5mg tablet |
| 1140860322 | pindolol+clopamide 10mg/5mg tablet |
| 1140860338 | viskaldix tablet |
| 1140860348 | atenixco 50mg/12.5mg tablet |
| 1140909706 | chlortalidone |
| 1141180772 | triamterene+chlortalidone 50mg/50mg tablet |
| 1141180778 | atenolol+chlortalidone |
| 1140860308 | metoprolol tartrate+chlorthalidone 100mg/12.5mg tablet |
| 1140923336 | co-tenidone |
| 1140864202 | chlorthalidone tablet+potassium m/r tablet 25mg/6.7mmol pack |
| 1140866144 | chlorthalidone |
| 1140851364 | hygroton k tablet combination pack |
| 1140866330 | triamterene+chlorthalidone 50mg/50mg tablet |
| 1140866410 | kalspare tablet |
| 1141146124 | atenolol+chlorthalidone |
| 1140866146 | hygroton 50mg tablet |
| 1140866084 | mefruside |
| 1140866086 | baycaron 25mg tablet |
| 1140866092 | metolazone |
| 1140866094 | metenix-5 tablet |
| 1140866096 | xuret 500micrograms tablet |
| 1140866108 | xipamide |
| 1140866110 | diurexan 20mg tablet |
| 1140866078 | indapamide |
| 1141180592 | perindopril+indapamide |
| 1140888922 | nindaxa 2.5mg tablet |
| 1141146378 | natrilix sr 1.5mg m/r tablet |
| 1140917068 | opumide 2.5mg tablet |
| 1141180598 | coversyl plus 4mg/1.25mg tablet |
| **NSAIDs** |  |
| 1140856214 | solprin 300mg dispersible tablet |
| 1140871168 | voltarol 25mg e/c tablet |
| 1140871100 | azapropazone |
| 1140853082 | ramodar 200mg tablet |
| 1140871450 | dysman-250 capsule |
| 1141176662 | celecoxib |
| 1140856312 | claradin 300mg tablet |
| 1140871180 | rhumalgan 25mg e/c tablet |
| 1140871102 | rheumox 300mg capsule |
| 1140853090 | ibular 200mg tablet |
| 1140871454 | contraflam 250mg capsule |
| 1141176668 | celebrex 100mg capsule |
| 1140856314 | laboprin 300mg tablet |
| 1140871188 | etodolac |
| 1140871578 | larapam 10mg capsule |
| 1140853094 | ibumetin 200mg tablet |
| 1140871542 | mefenamic acid |
| 1141176670 | celebrex 200mg capsule |
| 1140856332 | antoin dispersible tablet |
| 1140871196 | lodine 200mg tablet |
| 1140871582 | pirozip 10 capsule |
| 1140853100 | paxofen 200mg tablet |
| 1140871546 | ponstan 250mg capsule |
| 1141180126 | parecoxib |
| 1140856336 | codis dispersible tablet |
| 1140871248 | volraman 25mg e/c tablet |
| 1140871590 | flamatrol 10mg capsule |
| 1140871202 | fenbufen |
| 1140928840 | tolfenamic acid |
| 1141180140 | etoricoxib |
| 1140861800 | platet 100mg effervescent tablet |
| 1140871256 | valenac 25mg e/c tablet |
| 1140871654 | phenylbutazone product |
| 1140871206 | lederfen 300mg tablet |
| 1140928844 | clotam 200mg capsule |
| 1141180148 | arcoxia 60mg tablet |
| 1140861804 | angettes 75mg tablet |
| 1140871260 | diclozip-25 e/c tablet |
| 1140871660 | butacote 100mg e/c tablet |
| 1140871218 | fenbuzip 300mg tablet |
| 1141180150 | arcoxia 90mg tablet |
| 1140861806 | aspirin 75mg tablet |
| 1140871266 | arthrotec tablet |
| 1140871662 | butazone 100mg tablet |
| 1140871226 | fenprofen |
| 1141180152 | arcoxia 120mg tablet |
| 1140861808 | disprin cv 100mg m/r tablet |
| 1140871274 | isclofen 50mg e/c tablet |
| 1140871666 | piroxicam |
| 1140871228 | fenopron 300mg tablet |
| 1140864860 | nu-seals aspirin 75mg e/c tablet |
| 1140871276 | flamrase 25mg e/c tablet |
| 1140871672 | feldene 10mg capsule |
| 1140871236 | fluriprofen |
| 1140868226 | aspirin |
| 1140871336 | indomethacin |
| 1140875346 | tenoxicam |
| 1140871238 | froben 50mg tablet |
| 1140868258 | aspav dispersible tablet |
| 1140871344 | artracin 25mg capsule |
| 1140875350 | mobiflex 20mg tablet |
| 1140871310 | ibuprofen |
| 1140868282 | aspirin+methocarbamol 325mg/400mg tablet |
| 1140871348 | imbrilon 25mg capsule |
| 1140926732 | meloxicam |
| 1140871320 | arthrofen 200 tablet |
| 1140872040 | aspirin+metoclopramide 325mg/5mg effervescent tablet |
| 1140871354 | indocid 25mg capsule |
| 1140871370 | apsifen 200mg tablet |
| 1140882108 | aspirin+cyclizine hydrochloride 500mg/25mg tablet |
| 1140871360 | flexin-25 continus m/r tablet |
| 1140871374 | brufen 200mg tablet |
| 1140882190 | aspirin+glycine 500mg/133mg dispersible tablet |
| 1140871430 | rimacid 25mg capsule |
| 1140871386 | ebufac 200mg tablet |
| 1140882192 | disprin direct dispersible tablet |
| 1140871434 | indomod 25mg m/r capsule |
| 1140871388 | cuprofen 200mg tablet |
| 1140882268 | aspirin+codeine 300mg/8mg tablet |
| 1140871442 | indomax 25 capsule |
| 1140871392 | isisfen 400mg tablet |
| 1140882392 | aspirin+codeine |
| 1140871604 | sulindac |
| 1140871394 | fenbid 300mg spansule |
| 1140909480 | aspro clear maximum strength soluble tablet |
| 1140871606 | clinoril 100mg tablet |
| 1140871396 | lidifen 200mg tablet |
| 1140909772 | acetylsalicylic acid |
| 1140875268 | tolmetin |
| 1140871402 | codafen continus m/r tablet |
| 1140917408 | postmi 75mg dispersible tablet |
| 1140875270 | tolectin 200mg capsule |
| 1140871404 | junifen 100mg/5ml s/f suspension |
| 1140925942 | caprin 75mg e/c tablet |
| 1140875278 | acemetacin |
| 1140871406 | motrin 200mg tablet |
| 1141151924 | enprin 75mg e/c tablet |
| 1140884558 | ketorolac |
| 1140871408 | ibumed 400mg tablet |
| 1141157414 | salicylic acid product |
| 1140911560 | nambutenone |
| 1140871416 | rimafen 200mg tablet |
| 1141163138 | aspirin+papaveretum 500mg/7.71mg dispersible tablet |
| 1140921968 | lofensaid 25 tablet |
| 1140871462 | naproxen |
| 1141164044 | isosorbide mononitrate+aspirin |
| 1140925806 | aceclofenac |
| 1140871468 | laraflex 250mg tablet |
| 1141164050 | imazin xl 60mg/75mg m/r tablet |
| 1140925808 | preservex 100mg tablet |
| 1140871472 | naprosyn 250mg tablet |
| 1141167026 | caspac xl 162.5mg m/r capsule |
| 1140871482 | rheuflex-250 tablet |
| 1141167844 | dipyridamole+aspirin |
| 1140871484 | valrox 250mg tablet |
| 1141177826 | micropirin 75mg e/c tablet |
| 1140871490 | pranoxen continus 375mg m/r tablet |
| 1140871492 | arthrosin 250 tablet |
| 1140871506 | ketoprofen |
| 1140871514 | alrheumat 50mg capsule |
| 1140871516 | orudis 50mg capsule |
| 1140871522 | oruvail 100 m/r capsule |
| 1140871528 | ketovail 100mg m/r capsule |
| 1140871532 | ketonal 50mg capsule |
| 1140871556 | arthroxen 250mg tablet |
| 1140871564 | rimoxyn 250mg tablet |
| 1140871568 | nycopren 250mg e/c tablet |
| 1140871614 | tiaprofenic acid |
| 1140871616 | surgam 200mg tablet |
| 1140871628 | prosaid 250mg tablet |
| 1140871638 | napratec tablet combination pack |
| 1140878030 | ibuprofen+codeine phosphate |
| 1140910496 | propionic acid-ibuprofen |
| 1141153134 | anadin ibuprofen 200mg tablet |
| 1141157412 | ibuprofen product |
| 1141164746 | dexketoprofen |
| 1141164750 | keral 25mg tablet |
| 1141184546 | ibuprofen+pseudoephedrine hydrochloride |
| 1141187776 | nurofen 200mg |
| 1141190952 | cuprofen plus tablet |
| 1141194296 | lemsip flu 12hr ibuprofen+pseudoephedrine capsule |

Note: These medications' information was obtained from field 20003. ACEIs, angiotensin-converting enzyme inhibitors; ARBs, angiotensin II receptor blockers; BBs, *β*-blockers; CCBs, calcium channel blockers; NSAIDs, non-steroidal anti-inflammatory drugs.

Table S3. STROBE statement — checklist of items that should be included in reports of our study

| **Item No.** | **Section** | **Recommendation** | **Relevant text from the manuscript** |
| --- | --- | --- | --- |
|  | **Title and Abstract** | (a) Indicate the study's design with a commonly used term in the title or the abstract | Prospective cohort study. |
| 1 |  | (b) Provide in the abstract an informative and balanced summary of what was done and what was found | Prospective cohort study of 29,266 UK Biobank participants with pre-existing CVD but no PD at baseline. Participants were categorized by eGFR levels and followed for PD's incidence and all-cause mortality. Associations were assessed with multivariable Cox models adjusted for confounders. Higher baseline eGFR associated with lower PD risk (HR=0.77). Lower eGFR associated with progressively higher all-cause mortality risk. |
|  | **Introduction** |  |  |
| 2 | Background/  rationale | Explain the scientific background and rationale for the investigation being reported | Renal impairment is associated with increased risk of PD in the general population; however, the renal-PD link within CVD patients remains unclear given the high comorbidity of renal dysfunction and elevated PD risk among this special population. |
| 3 | Objectives | State specific objectives, including any prespecified hypotheses | Objective: To establish the relationship between eGFR and PD risk and all-cause mortality in patients with pre-existing CVD.  Hypothesis: eGFR decline is associated with higher PD incidence and mortality in this population. |
|  | **Methods** |  |  |
| 4 | Study design | Present key elements of study design early in the paper | Figure 1. Roadmap of the study. |
| 5 | Setting | Describe the setting, locations, and relevant dates, including periods of recruitment, exposure, follow-up, and data collection | The study was set in the UK Biobank, a prospective cohort recruiting 500,000 participants aged 37-73 from 22 centers in England, Wales, and Scotland during 2006-2010. The analysis included 29,266 CVD patients of European ancestry with baseline eGFRcr and eGFRcys measurements. Follow-up ran from baseline to PD diagnosis, death, or July 19, 2022 (median 13.1 years). Baseline data covered sociodemographic, lifestyle, health status, and biological samples. |
| 6 | Participants | (a) Give the eligibility criteria, and the sources and methods of selection of participants. Describe methods of follow-up | The eligibility criteria included participants of European ancestry from the UK Biobank, aged 37-73 years at recruitment (2006-2010), with baseline CVD, and available serum creatinine and cystatin C measurements. Exclusions were those with missing renal function data, loss to follow-up, or withdrawal of consent. Participants were selected from 22 assessment centers across England, Wales, and Scotland. Follow-up was conducted via electronic health records and national death registries, tracking incident PD (ICD-10 code) and all-cause mortality from baseline to PD diagnosis, death, or July 19, 2022. |
|  |  | (b) For matched studies, give matching criteria and number of exposed and unexposed | In the nested case-control analysis, incident PD cases and decedents were matched 1:1 to controls using baseline covariates (sociodemographic, lifestyle factors, health status, medication use, and genetic principal components). For PD, 489 cases were matched to 489 controls; for all-cause mortality, 5,919 decedents were matched to 5,919 surviving controls. Matching ensured covariate balance between PD and non-PD groups to assess pre-outcome eGFR trajectories over 15 years preceding PD diagnosis or death. |
| 7 | Variables | Clearly define all outcomes, exposures, predictors, potential confounders, and effect modifiers. Give diagnostic criteria, if applicable | Exposures were renal function metrics: eGFRcr and eGFRcys, with lower values indicating impairment. Outcomes were incident PD (defined by ICD-10 code and neurologist validation) and all-cause mortality. We selected the following baseline covariates: socioeconomic factors (TDI, educational level, and income), behavioral factors (alcohol consumption status, smoking status, physical activity status, and healthy diet score), health status (BMI, type 2 diabetes, hypertension, dyslipidemia), top-frequency CVD medication use (ACEIs, ARBs, BBs, CCBs, diuretics, NSAIDs), family history of PD and top ten genetic principal components. |
| 8 | Data sources/ measurement | For each variable of interest, give sources of data and details of methods of assessment (measurement). Describe comparability of assessment methods if there is more than one group | For renal function, eGFRcr and eGFRcys were calculated using CKD-EPI equations from serum creatinine and cystatin C measurements at baseline. PD was identified via ICD-10 codes in electronic health records, validated by neurologists. All-cause mortality data came from national death registries. Covariates (sociodemographic, lifestyle, health status, medications) were from baseline questionnaires, physical exams, and biological samples. Genetic principal components were derived from genotyping. Assessment methods were consistent across participants, with missing covariate values imputed using multiple chained equations, ensuring comparability. |
| 9 | Bias | Describe any efforts to address potential sources of bias | To address potential bias, the study employed multiple strategies. Reverse causality was mitigated by excluding events within the first 2 years of follow-up. Confounding was minimized using GPS, IPW and E-value to balance covariates. Cause-specific hazards regression adjusted for competing risks of non-PD deaths. Restricted cubic splines evaluated nonlinear associations, and time-stratified analyses assessed temporal heterogeneity. Multiple imputation handled missing data, ensuring robust results. |
| 10 | Study size | Explain how the study size was arrived at | The study included 29,266 CVD patients from the UK Biobank. Eligibility required European ancestry, baseline CVD, available eGFRcr/eGFRcys, and no loss to follow-up/consent withdrawal. Sample size ensured statistical power for multivariable analyses of renal function's association with PD and mortality. |
| 11 | Quantitative variables | Explain how quantitative variables were handled in the analyses. If applicable, describe which groupings were chosen and why | Quantitative variables like eGFRcr and eGFRcys were analyzed both continuously and categorically. Continuously, eGFR was standardized to improve model interpretability. Categorically, they were grouped into normal (≥90 ml/min/1.73m^2^), mildly reduced (60-90 ml/min/1.73m^2^), and CKD (<60 ml/min/1.73m^2^) based on clinical guidelines to assess risk gradients. This dual approach balanced statistical flexibility with clinical relevance, enabling dose-response evaluation and stratification for risk prediction. |
| 12 | Statistical methods | (a) Describe all statistical methods, including those used to control for confounding | This study analyzed UK Biobank CVD patients using multivariable Cox proportional hazards regression to assess associations between baseline eGFR (continuous and categorized) and incident PD or all-cause mortality, adjusting for socioeconomic, behavioral, health status, medication use, and genetic covariates. Restricted cubic splines modeled non-linear exposure-outcome relationships. Controls for confounding included: (1) exclusion of early outcome cases to reduce reverse causation; (2) GPS analysis with IPW to balance covariates across eGFR; (3) E-values to quantify susceptibility to residual confounding; and (4) cause-specific hazards models accounting for competing mortality risk. A nested case-control analysis used 1:1 Propensity Score Matching (PSM) to map longitudinal pre-PD/pre-death eGFR trajectories over 14 years. For prediction, the PREDICT-PD algorithm was validated and enhanced with CKD status using internal validation; performance gains were assessed with Monte Carlo cross-validation and machine learning models. Multiple imputation handled missing covariate data. |
|  |  | (b) Describe any methods used to examine subgroups and interactions | Null. |
|  |  | (c) Explain how missing data were addressed | The method of multivariate imputations with chained equations was implemented to impute missing values of each covariate. There is no missing value for exposures and outcomes. |
|  |  | (d) If applicable, explain how loss to follow-up was addressed | We excluded participants who withdrawn from the survey. |
|  |  | (e) Describe any sensitivity analyses | The study conducted several sensitivity analyses: excluding events within the first 2 years to address reverse causality, using GPS, IPW and E-value to balance confounders, applying cause-specific hazards regression for PD by censoring non-PD deaths. These validated the robustness of associations between renal function and risks of PD and all-cause mortality. |
|  | **Results** |  |  |
| 13 | Participants | (a) Report numbers of individuals at each stage of study—eg numbers potentially eligible, examined for eligibility, confirmed eligible, included in the study, completing follow-up, and analysed | The UK Biobank initially recruited 500,000 participants. The study included 29,266 participants of European ancestry with baseline CVD, excluding those with missing renal function data, loss to follow-up, or consent withdrawal. All included participants were analyzed, with a median follow-up of 13.1 years (IQR 12.2-14.0 yrs), tracking 489 incident PD cases and 5,919 all-cause deaths. |
|  |  | (b) Give reasons for non-participation at each stage | Non-participation reasons included missing serum creatinine or cystatin C measurements, loss to follow-up during the study, and withdrawal of informed consent. Participants without baseline CVD or of non-European ancestry were also excluded to ensure population homogeneity and statistical power for the analysis of renal function in CVD patients. |
|  |  | (c) Consider use of a flow diagram | We provided the flow diagram in Figure 1. |
| 14 | Descriptive data | (a) Give characteristics of study participants (eg demographic, clinical, social) and information on exposures and potential confounders | The study involved 29,266 CVD patients (33.4% female, mean age 62.2 yrs). Demographically, they were of European ancestry, with 80.1% having no college education and mean TDI -0.77. Clinically, mean eGFRcr was 88.2 mL/min/1.73m², eGFRcys 78.3 mL/min/1.73m²; 11.1% had type 2 diabetes. Socially, 15.8% were former smokers, 5.7% former drinkers. |
|  |  | (b) Indicate number of participants with missing data for each variable of interest | For household income, 4,922 (16.82%) had missing data. For physical activity, 6,322 (21.60%) were missing. Health diet score had 4,572 (15.62%) missing values. TDI had 31 (0.11%) missing, and educational qualifications had 1 (0.00%) missing. Other variables like BMI, smoking, alcohol use, and comorbidities had minimal or no missing data, handled via multiple imputation. |
|  |  | (c) Cohort study—Summarize follow-up time (e.g., average and total amount) | The median follow-up time was 13.1 years (interquartile range: 12.2-14.0 years). |
| 15 | Outcome data | Cohort study—Report numbers of outcome events or summary measures over time;  Case-control study—Report numbers in each exposure category, or summary measures of exposure;  Cross-sectional study—Report numbers of outcome events or summary measures | During follow-up (median 13.1 years), there were 489 incident PD cases and 5,919 all-cause deaths. |
| 16 | Main results | (a) Give unadjusted estimates and, if applicable, confounder-adjusted estimates and their precision (e.g., 95% confidence interval). Make clear which confounders were adjusted for and why they were included | In the study, multivariable-adjusted Cox regression models were used to assess the associations between renal function and risks of incident PD and all-cause mortality. For PD risk, each 1-SD decrease in eGFRcr was associated with a 12.7% (4.7%~20.0%) increase, and each 1-SD decrease in eGFRcys was linked to a 10.1% (1.2%~18.2%) elevation. Compared to normal renal function, CKD defined by eGFRcr and eGFRcys was associated with a 60.1% (12.0%~128.9%) and 38.4% (3.0%~85.9%) higher PD risk, respectively. For all-cause mortality, each 1-SD decrease in eGFRcr and eGFRcys was associated with a 22.8% (21.1%~24.6%) and a significant inverse association (HR=0.641 [0.625~0.657]), respectively. CKD defined by eGFRcr and eGFRcys was associated with a 158.9% (137.7%~182.0%) and 233.5% (207.6%~261.7%) higher mortality risk, respectively. The models were adjusted for socioeconomic factors (TDI, educational level, and income), behavioral factors (alcohol consumption status, smoking status, physical activity status, and healthy diet score), health status (BMI, type 2 diabetes, hypertension, dyslipidemia), top-frequency CVD medication use (ACEIs, ARBs, BBs, CCBs, diuretics, NSAIDs), family history of PD and top ten genetic principal components. These confounders were included as they are potential risk factors or correlates of both renal function and the outcomes, helping to mitigate confounding and ensure the validity of the associations. |
|  |  | (b) Report category boundaries when continuous variables were categorized | When continuous variables of renal function were categorized, the study used clinical guidelines to define three groups for both eGFRcr and eGFRcys. Normal renal function was set as eGFR≥90 ml/min/1.73m², mildly reduced renal function as 60≤eGFR<90 ml/min/1.73m², and CKD as eGFR<60 ml/min/1.73m². This categorization aligns with standard clinical definitions to facilitate interpretation of renal dysfunction severity and its associations with PD and mortality. |
|  |  | (c) If relevant, consider translating estimates of relative risk into absolute risk for a meaningful time period | Null. |
| 17 | Other analyses | Report other analyses done—e.g., analyses of subgroups and interactions, and sensitivity analyses | The study performed sensitivity analyses including a 2-year lag period exclusion (minimal HR variation), GPS adjustment (e.g., eGFRcr-based CKD mortality HR=2.85 [2.62~3.10]), IPW weighting (eGFRcys-CKD mortality HR=3.97 [3.67~4.30]), and cause-specific hazards regression (PD risk independent of mortality). Time-stratified analysis showed eGFRcr-CKD associated with PD only in intermediate phases (HR=2.26 [1.39~3.68]), while mortality associations persisted. |
|  | **Discussion** |  |  |
| 18 | Key results | Summarise key results with reference to study objectives | The significant inverse associations observed for both eGFR metrics demonstrate that reduced renal function is an independent risk factor for PD and all-cause mortality in this high-risk population. Furthermore, our longitudinal trajectory analyses revealed that a slow, progressive decline in eGFR begins over 14 years prior to PD diagnosis, establishing renal dysfunction as a protracted, prodromal process rather than a mere consequence of established disease. Critically, incorporating eGFR-defined CKD status significantly enhanced the discrimination and prediction accuracy of the PREDICT-PD algorithm in this high-risk population using routine clinical data. |
| 19 | Limitations | Discuss limitations of the study, considering sources of potential bias or imprecision. Discuss both direction and magnitude of any potential bias | Our findings must be interpreted within the context of the study's limitations. First, the restriction to individuals of European ancestry ensures population homogeneity but limits generalizability to other ethnic groups known to have different CKD prevalence and PD risks. Second, serum creatinine and cystatin C levels, while widely used to estimate GFR, do not completely equate to measured GF. Although we have comprehensively examined their associations with PD risk and mortality at the phenotypic level, causal inferences remain limited; further mechanistic investigations and intervention trials are warranted to establish causality. Third, our prediction enhancement was implemented within the specific PREDICT-PD algorithm; the magnitude of improvement might differ if applied to other prediction models. Finally, this study focused on prevalent CVD patients, so results may not directly apply to healthy populations or those without CVD. |
| 20 | Interpretation | Give a cautious overall interpretation of results considering objectives, limitations, multiplicity of analyses, results from similar studies, and other relevant evidence | This study shows that reduced renal function, measured by eGFRcr and eGFRcys, is independently associated with higher risks of PD and all-cause mortality in CVD patients, with CKD conferring the greatest risks. The dose-dependent relationships and distinct temporal trajectories (eGFR decline starting >14 years pre-PD, accelerating pre-mortality) are consistent across sensitivity analyses, mitigating confounding and reverse causality. While limitations like single baseline eGFR assessment, European ancestry bias, and residual genetic confounding exist, findings align with prior studies linking renal dysfunction to PD and mortality. The dual-biomarker approach enhances risk stratification, supporting routine renal function monitoring in CVD populations to inform early neuroprotective and cardiorenal interventions, though broader validation in diverse cohorts is needed. |
| 21 | Generalizability | Discuss the generalizability (external validity) of the study results | The study's generalizability is limited by its reliance on a UK Biobank cohort predominantly of European ancestry, which may not reflect ethnic/racial differences in renal function-PD/mortality associations. Additionally, the focus on CVD patients with baseline eGFR measurements may not apply to general populations or those with different comorbidity profiles. The use of baseline eGFR without longitudinal monitoring might not capture dynamic renal function changes, potentially affecting external validity. Findings require validation in more diverse cohorts with varied ethnic backgrounds and comorbidity patterns. |
|  | **Other information** |  |  |
| 22 | Funding | Give the source of funding and the role of the funders for the present study and, if applicable, for the original study on which the present article is based | The research of Ping Zeng was supported in part by the Youth Foundation of Humanity and Social Science funded by Ministry of Education of China (18YJC910002). The research of Chu Zheng was supported by the Project of Philosophy and Social Science Research in Colleges and Universities of Jiangsu Province (2024SJYB0809). |

Note: Information on the STROBE Initiative is available at https://www.strobe-statement.org [4]. CVD, cardiovascular disease; CKD, chronic kidney disease; PD, Parkinson's disease; eGFR, estimated glomerular filtration rate; eGFRcr, estimated glomerular filtration rate based on creatinine; eGFRcys, estimated glomerular filtration rate based on cystatin C; BMI, body mass index; TDI, Townsend deprivation index; ACEIs, angiotensin-converting enzyme inhibitors; ARBs, angiotensin II receptor blockers; BBs, *β*-blockers; CCBs, calcium channel blockers; NSAIDs, non-steroidal anti-inflammatory drugs; SD, standard deviation; IQR, interquartile range; HR, hazard ratio; CI, confidence interval; GPS, generalized propensity scores; IPW, inverse probability weights.

Table S4. Phenotypic association of eGFR with PD in sensitivity analysis.

| **Outcome** | **Exposure** | **2-year lag analyses** | |  | **Confounding adjustment analyses** | | | | |  | **Cause-specific hazards regression** | |
| --- | --- | --- | --- | --- | --- | --- | --- | --- | --- | --- | --- | --- |
|  |  |  |  |  | **GPS** | | **IPW** | | **E-value** |  |  |  |
|  |  | **HRs (95%CIs)** | ***P*** |  | **HRs (95%CIs)** | ***P*** | **HRs (95%CIs)** | ***P*** | **E-values (95%CIs)** |  | **HRs (95%CI)** | ***P*** |
| PD | eGFRcr | 0.869 (0.794~0.950) | 2.08×10^-3^ |  | 0.875 (0.802~0.956) | 3.07×10^-3^ | 0.855 (0.787~0.929) | 2.21×10^-4^ | 1.613 (1.855~1.362) |  | 0.851 (0.783~0.924) | 1.22×10^-4^ |
|  | G1 | Ref. |  |  | Ref. |  | Ref. |  | Ref. |  | Ref. |  |
|  | G2 | 1.013 (0.836~1.228) | 0.895 |  | 0.994 (0.824~1.199) | 0.949 | 1.023 (0.849~1.233) | 0.807 | N/A |  | 1.032 (0.855~1.246) | 0.741 |
|  | G3 | 1.604 (1.111~2.317) | 0.012 |  | 1.616 (1.131~2.308) | 8.33×10^-3^ | 1.742 (1.223~2.481) | 2.09×10^-3^ | 2.880 (1.746~4.398) |  | 1.872 (1.310~2.674) | 5.76×10^-4^ |
|  | eGFRcys | 0.890 (0.808~0.980) | 0.018 |  | 0.903 (0.821~0.993) | 0.036 | 0.903 (0.823~0.991) | 0.031 | 1.452 (1.725~1.108) |  | 0.861 (0.787~0.943) | 1.16×10^-3^ |
|  | G1 | Ref. |  |  | Ref. |  | Ref. |  | Ref. |  | Ref. |  |
|  | G2 | 1.084 (0.872~1.348) | 0.469 |  | 1.058 (0.855~1.309) | 0.606 | 1.084 (0.878~1.339) | 0.453 | N/A |  | 1.100 (0.889~1.361) | 0.380 |
|  | G3 | 1.398 (1.033~1.893) | 0.030 |  | 1.372 (1.020~1.844) | 0.037 | 1.428 (1.072~1.901) | 0.015 | 2.209 (1.35~3.209) |  | 1.610 (1.199~2.162) | 1.53×10^-3^ |
| All-cause mortality | eGFRcr | 0.769 (0.751~0.787) | 1.22×10^-108^ |  | 0.762 (0.745~0.780) | 1.73×10^-114^ | 0.762 (0.743~0.780) | 1.21×10^-105^ | 1.954 (2.027~1.882) |  |  |  |
|  | G1 | Ref. |  |  | Ref. |  | Ref. |  |  |  |  |  |
|  | G2 | 1.248 (1.179~1.321) | 2.15×10^-14^ |  | 1.151 (1.090~1.215) | 4.24×10^-7^ | 1.154 (1.093~1.218) | 2.04×10^-7^ | 1.576 (1.413~1.734) |  |  |  |
|  | G3 | 2.606 (2.382~2.852) | 8.38×10^-97^ |  | 2.848 (2.616~3.102) | 4.23×10^-128^ | 2.909 (2.673~3.166) | 5.12×10^-135^ | 5.266 (4.788~5.785) |  |  |  |
|  | eGFRcys | 0.646 (0.629~0.663) | 1.68×10^-236^ |  | 0.632 (0.616~0.648) | 1.64×10^-267^ | 0.603 (0.588~0.619) | 0.000 | 2.703 (2.795~2.614) |  |  |  |
|  | G1 | Ref. |  |  | Ref. |  | Ref. |  |  |  |  |  |
|  | G2 | 1.514 (1.405~1.632) | 1.86×10^-27^ |  | 1.439 (1.339~1.546) | 3.69×10^-23^ | 1.553 (1.446~1.669) | 1.65×10^-33^ | 2.481 (2.249~2.725) |  |  |  |
|  | G3 | 3.310 (3.041~3.603) | 9.82×10^-169^ |  | 3.332 (3.071~3.616) | 2.59×10^-183^ | 3.972 (3.670~4.299) | 4.91×10^-256^ | 7.408 (6.800~8.065) |  |  |  |

Note: G1, normal renal function group; G2, mildly reduced renal function group; G3, CKD group; PD, Parkinson's disease; eGFRcr, estimated glomerular filtration rate based on creatinine; eGFRcys, estimated glomerular filtration rate based on cystatin C; CKD, chronic kidney disease; HRs, hazard ratios; CIs, confidence intervals; Ref., reference group.

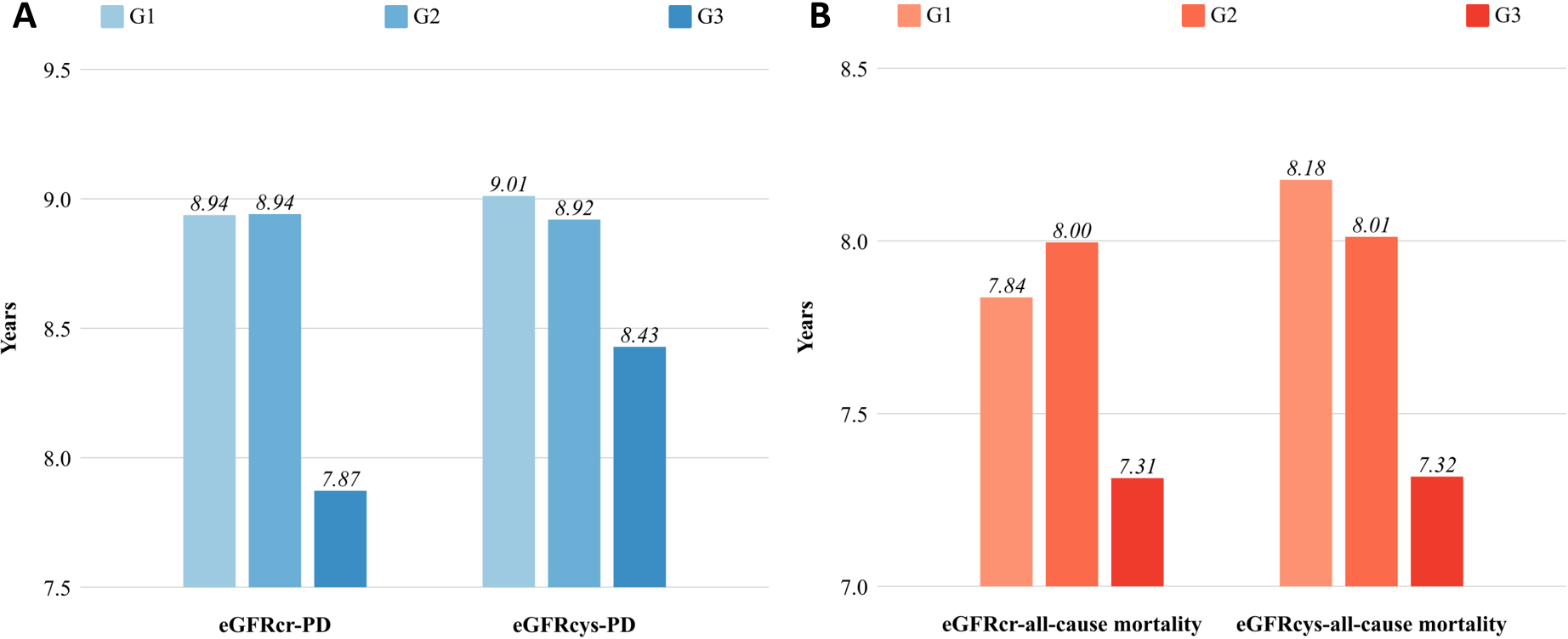

Figure S1. (A) Mean time to PD onset across three renal function categories. (B) Mean survival time across three renal function categories. G1, normal renal function group; G2, mildly reduced renal function group; G3, CKD group; PD, Parkinson's disease; eGFRcr, estimated glomerular filtration rate based on creatinine; eGFRcys, estimated glomerular filtration rate based on cystatin C; CKD, chronic kidney disease; HR, hazard ratio; CI, confidence interval.

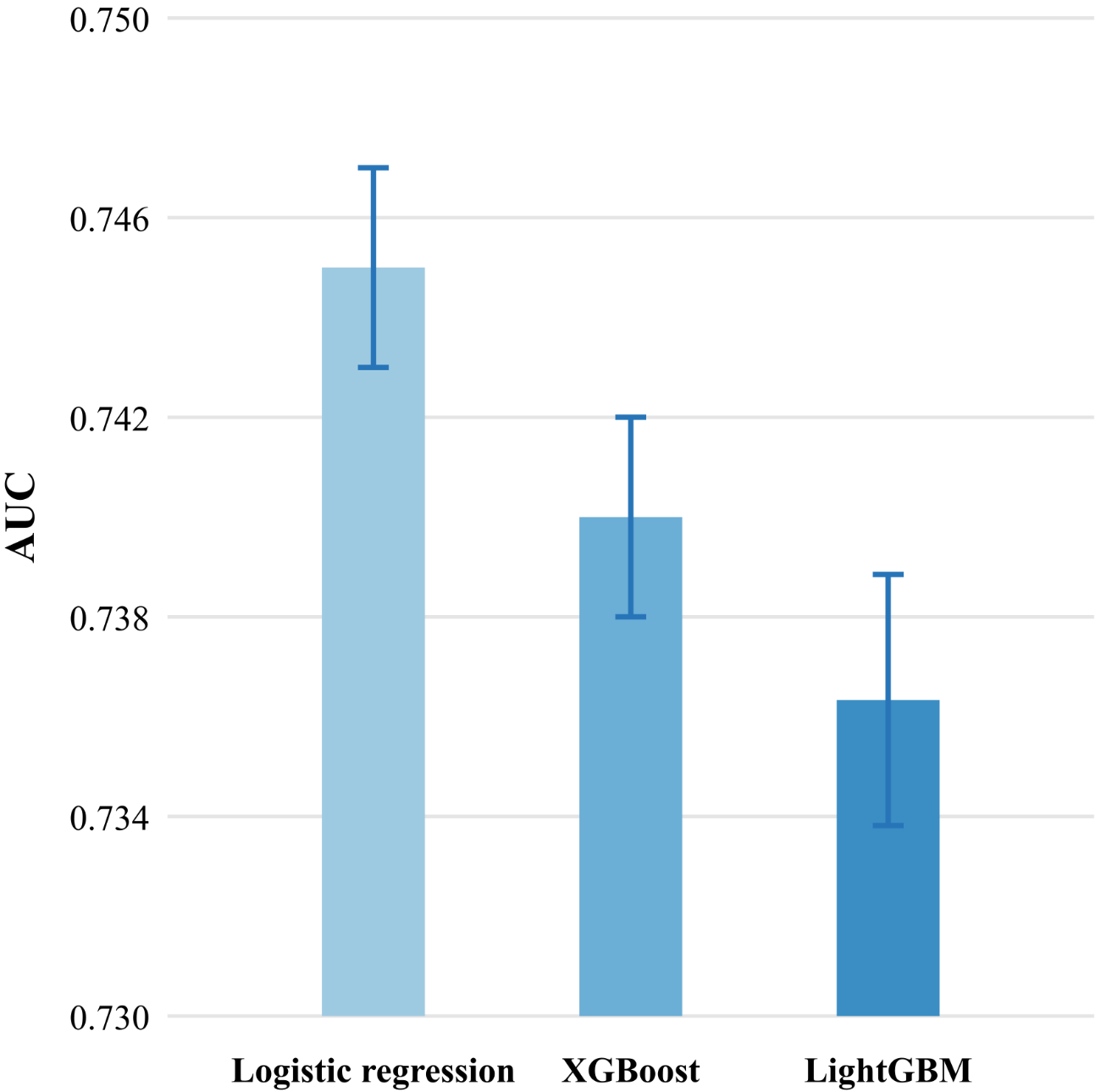

Figure S2. The performance of predicting the incident PD using the variable *P*_0_ through three methods. *P*_0_, original PD probability; AUC, area under the receiver operating characteristic curve.
